# The impact of COVID-19 on gastrointestinal diseases in vaccinated and unvaccinated individuals: a population-based cohort study in England

**DOI:** 10.64898/2025.12.01.25341357

**Authors:** Marwa Al Arab, Genevieve Cezard, Colin Crooks, Yinghui Wei, Elsie Horne, Tom Palmer, Jose I Cuitun Coronado, Alex Walker, Millie Green, Louis Fisher, Jon Massey, Simon Davy, Amir Mehrkar, Seb Bacon, Ben Goldacre, Nishi Chaturvedi, Angela Wood, Jonathan A C Sterne, Venexia Walker, Rachel Denholm

## Abstract

**Background:** COVID-19 is associated with higher rates of gastrointestinal diseases, though the duration of this effect, and the role of vaccination and COVID-19 severity remain uncertain.

**Objective:** To investigate the relationship between COVID-19 and gastrointestinal diseases, by vaccination status in hospitalised and non-hospitalised patients.

**Design:** With NHS England approval, OpenSAFELY-TPP was used to access linked data from 24 million English adults. We defined three cohorts: “Pre-vaccination” (January 2020-June 2021), “vaccinated” and “unvaccinated” (June-December 2021). We estimated adjusted hazard ratios (aHRs) comparing the incidence of 10 gastrointestinal diseases, including upper and lower gastrointestinal bleeding, after versus before or without a COVID-19 diagnosis overall and by COVID-19 severity.

**Results:** COVID-19 diagnosis was associated with elevated incidence of gastrointestinal diseases, particularly after hospitalised disease. In the pre-vaccination cohort (n=18,422,781), the adjusted hazard ratios (aHRs) for upper gastrointestinal bleeding after hospitalised COVID-19 were 20.1 (95% CI 18.4–22.1) in weeks 1–4 and 1.90 (1.63–2.22) in weeks 53–102. The corresponding aHRs after non-hospitalised COVID-19 were 1.92 (1.74–2.11) and 1.34 (1.23– 1.45) respectively. Across time periods, aHRs for gastrointestinal diseases were lower in vaccinated (n=14,948,727) than in unvaccinated (n=3,479,043) individuals. These patterns were similar across gastrointestinal diseases.

**Conclusion:** The incidence of gastrointestinal disease is elevated for up to two years among individuals hospitalised following a COVID-19 diagnosis. Increases in incidence are attenuated among vaccinated individuals. Monitoring for gastrointestinal diseases after severe COVID-19 and promotion of vaccination in vulnerable groups are key to reducing long-term burden.

**KEY MESSAGES:** *What is already known on this topic:*

- Acute symptoms of COVID-19 include gastrointestinal problems, and there is existing evidence that those with a COVID-19 diagnosis are more likely to experience long-term gastrointestinal complications, especially if they were hospitalised.
- Previous studies were either limited in size, lacked representativeness of the population, or had short follow-up (e.g. during hospitalisation). The role of vaccination, COVID-19 severity or peoples’ characteristics (e.g. age or sex) were not considered.

*What this study adds:*

- Based on population-wide data on 18.4 million individuals, rates of 10 gastrointestinal diseases are elevated after COVID-19, compared with rates before or without COVID-19 diagnosis.
- The elevation in rates of gastrointestinal diseases after COVID-19 diagnosis was less marked in people who were vaccinated before COVID-19 diagnosis.
- The elevation in rates was markedly higher, and persisted for up to two years, among individuals hospitalised with COVID-19, compared with those who were not hospitalised.

*How this study might affect research, practice or policy:*

- We show for the first time that the incidence of both acute and chronic gastrointestinal diseases remains elevated for up to two years after severe COVID-19 leading to hospitalisation.
- Our findings highlight the protective role of COVID-19 vaccination in reducing severe COVID-19 and its subsequent long-term gastrointestinal complications, reinforcing the importance of vaccination as a public health measure.
- Individuals who had severe COVID-19, regardless of vaccination status, should be prioritised for clinical monitoring for gastrointestinal diseases.

## INTRODUCTION

COVID-19 can lead to long-term adverse outcomes that affect several body systems, including the gastrointestinal system. [1,2] Potential mechanisms include direct infection of gastrointestinal epithelial cells via ACE2 receptors [3], immune dysregulation, persistent viral presence in the gut or microvascular injury. [4,5] Previous studies have identified associations of COVID-19 with gastrointestinal diseases in both hospitalised [6–12] and non-hospitalised individuals [13–17], for up to 2.6 years after infection. [18,19] A recent meta-analysis estimated that up to 22% of long COVID patients experience gastrointestinal symptoms, including dyspepsia, abdominal pain, loss of appetite, and irritable bowel syndrome (IBS). [20] However, published studies are limited by single-centre data, short follow-up time, or non-representative samples, and many focused only on symptoms or a narrow range of diseases.

COVID-19 vaccination substantially reduces the risk of severe COVID-19 [21], but there is mixed evidence on its protective effect against post-acute sequelae. Some studies found that vaccination reduced the risk of post-acute sequelae of COVID-19 including neurological and gastrointestinal symptoms, such as abdominal pain, loss of appetite, constipation and diarrhoea [22]. Other studies found no protection by COVID-19 vaccination against post-acute sequelae, such as anxiety, sleep disorders, and type 2 diabetes, particularly in older individuals. [23] Evidence on the impact of vaccination on post-acute gastrointestinal diseases following COVID-19 remains limited.

Using linked electronic health records from England, we compared rates of 10 acute and chronic gastrointestinal diseases after a COVID-19 diagnosis with rates before or without COVID-19 across three cohorts defined by vaccination status (pre-vaccination, vaccinated and unvaccinated). We also examined associations in subgroups defined by COVID-19 severity, age, sex, ethnicity and prior gastrointestinal history.

## METHODS

### Study design and data sources

We analysed data available through the OpenSAFELY platform, which provides secure, privacy-protecting access to linked electronic health records for 24 million people registered with English general practices (GPs) using the TPP SystmOne software – approximately 40% of the English population. Primary care data was linked via pseudonymised NHS number to Secondary Uses Service (SUS) secondary care data, Office of National Statistics (ONS) Death Registry, Second Generation Surveillance System (SGSS) COVID-19 testing data and the Index of Multiple Deprivation (IMD). COVID-19 vaccination records (National Immunisation Management System) are available within TPP primary care data.

The gastrointestinal diseases studied were: upper and lower gastrointestinal bleeding, acute pancreatitis, appendicitis, gastro-oesophageal reflux, dyspepsia, peptic ulcer, non-alcoholic steatohepatitis, gallstones and irritable bowel syndrome (IBS). Outcomes were defined using pre-specified codelists developed for this study (https://github.com/opensafely/post-covid-gastrointestinal/tree/main/codelists). Date of outcome was defined as the earliest of: a relevant SNOMED CT code recorded in primary care; the start of a secondary care episode with a corresponding ICD-10 diagnosis code in any position; or death with an ICD-10 code indicating the diagnosis as either a primary or underlying cause.

Date of COVID-19 diagnosis was defined as the earliest of: positive SARS-CoV-2 PCR or antigen test recorded in SGSS; confirmed COVID-19 diagnosis recorded in primary care; start of an episode with a confirmed diagnosis in any position for a hospital admission record in SUS; or death with COVID-19 listed as primary or underlying cause of death in the ONS registry. People with a hospital admission record in SUS that included a confirmed COVID-19 in the primary position and within 28 days of first COVID-19 diagnosis were defined as having had ‘hospitalised COVID-19’. All other COVID-19 diagnoses were classified as ‘non-hospitalised COVID-19’. COVID-19 hospitalisation was used as a proxy for disease severity, with hospitalised cases representing more severe disease and non-hospitalised cases representing less severe disease.

Covariates identified as potential confounders at baseline included age, sex, ethnicity, area socioeconomic deprivation, smoking status, obesity, alcohol consumption above the recommended limit, care home residence, health care worker, and number of GP-patient interactions in 2019. Binary indicators for history of comorbidities (including prior symptoms and gastrointestinal diseases), H. pylori infection, gastrointestinal surgical procedures, and medication use (NSAIDs, aspirin, anticoagulants, and antidepressants) were also included (**Supplementary Table 1**).

### Study population

We defined three cohorts based on vaccination status and predominant SARS-CoV-2 variant (**Supplementary Table 2; Supplementary Figure 1**). In the ‘pre-vaccination’ cohort, when Alpha variant was dominant, exposure was defined as a COVID-19 diagnosis recorded between January 1, 2020 (baseline), and the earliest of eligibility for COVID-19 vaccination, date of first vaccination, or June 18, 2021 (when all adults became eligible for vaccination). Outcomes were ascertained from baseline until the earliest of: outcome event date, date of death, or December 14, 2021. In the ‘vaccinated’ and ‘unvaccinated’ cohorts, the follow-up period was defined between June 1, 2021 (baseline), when the Delta variant was dominant, and December 14, 2021, when Omicron became the dominant variant.[25] In the ‘vaccinated’ cohort, follow-up started at the later of baseline or two weeks after a second COVID-19 vaccination and ended at the earliest of the: outcome event date, date of death or December 14, 2021. In the ‘unvaccinated’ cohort, individuals were included if they had not received a COVID-19 vaccine within 12 weeks after becoming eligible. Follow-up started at the later of baseline or 12 weeks after vaccination eligibility and ended at the earliest of: outcome event date, date of death, date of first vaccination, or December 14, 2021.

In each cohort, individuals were eligible if they were alive with a known age (between 18 and 110), sex, deprivation, and region at baseline, and were registered with a GP in England for at least 6 months before the study start date. Individuals were excluded if they had any record of COVID-19 diagnosis before baseline. In the vaccinated cohort, individuals were excluded if they received a vaccination before the start of the vaccine rollout on December 8th 2020 (indicating an error or participation in a randomized trial); their second dose was dated before their first dose vaccination (contradictory vaccine record); their second dose was less than three weeks after their first dose; or they received mixed vaccine brands before this was permitted on May 7, 2021. In the unvaccinated cohort, individuals were excluded if they had any record of a COVID-19 vaccination before June 1, 2021. Vaccine eligibility was defined using the Joint Committee on Vaccination and Immunisation (JCVI) groupings. Individuals who could not be assigned to a JCVI group were excluded. We also excluded individuals with a history of Crohn’s disease, Coeliac disease or cirrhosis, and for the appendicitis analyses, those with prior diagnoses of appendicitis.

### Statistical analyses

Baseline demographics and clinical characteristics were summarised for each cohort. For each outcome, we calculated the number of events, person-years follow-up and incidence rates per 100,000 person-years before and after COVID-19, separately for individuals with hospitalised, non-hospitalised COVID-19 and no COVID-19 across each cohort.

For each outcome, we conducted time to first event analyses using Cox models with calendar timescale, with the cohort-specific baseline as the origin. We estimated hazard ratios (HRs) comparing follow-up after to follow-up before or without COVID-19. Follow-up after COVID-19 was split into the day of COVID-19 diagnosis (‘day 0’), the remainder of 1-4 weeks, and 5-28 weeks after COVID-19 diagnosis for all cohorts, and additionally 29-52 and 53-102 weeks after COVID-19 diagnosis for the pre-vaccination cohort. We focused inferences on events after day 0, because the ordering of exposure and outcome events on day 0 cannot be determined.

For computational efficiency, we included all individuals with the outcome event or exposure, and a 20% random sample of those without either. We used inverse probability weights to adjust for the sampling and used robust standard errors to derive confidence intervals.

For each outcome across the three cohorts, we reported both age- and sex-adjusted and maximally adjusted (for all potential confounders) HRs. Age was modelled using restricted cubic splines unless otherwise specified. All models were stratified by region to account for variation in baseline hazard across regions. Subgroup analyses according to age, sex, ethnicity, pre-existing gastrointestinal diseases, and prior gastrointestinal surgical procedures were conducted.

We calculated absolute excess risk 28 weeks after COVID-19 diagnosis, excluding events recorded on the day of COVID-19 diagnosis (‘day 0’) and weighted by the proportion of people in age strata in the pre-vaccination cohort. Additional details are provided in the supplementary methods.

### Data sharing

Analyses were performed according to a pre-specified protocol which is publicly available along with code lists and analysis code at https://github.com/opensafely/post-covid-gastrointestinal. This study was approved by the Health Research Authority [REC reference 22/PR/0095] and by the University of Bristol’s Faculty of Health Sciences Ethics Committee [reference 117269] (see “Information governance and ethical approval”).

All data were linked, stored and analysed securely using the OpenSAFELY platform, https://www.opensafely.org/, as part of the NHS England OpenSAFELY COVID-19 service. Data include pseudonymised data such as coded diagnoses, medications and physiological parameters. All code is shared openly for review and re-use under MIT open license https://github.com/opensafely/post-covid-gastrointestinal. Detailed pseudonymised patient data is potentially re-identifiable and therefore not shared.

Data management and analyses were conducted in Python version 3.8.10, R version 4.0.2 and Stata/MP version 16.1.

## RESULTS

Outcomes are presented based on acuity, starting with acute events associated with hospital admissions (upper gastrointestinal bleeding, lower gastrointestinal bleeding, acute pancreatitis and appendicitis), followed by more chronic diseases (gastro-oesophageal reflux, dyspepsia, peptic ulcer, nonalcoholic steatohepatits, gallstones and irritable bowel syndrome). Outcomes with insufficient event counts to run models in all cohorts, and by hospitalised COVID-19 status, are included in the supplementary material only, with findings briefly summarised in the text.

### Cohort characteristics

The pre-vaccination cohort included 18,422,781 people of whom 1,000,437 (5.43%) received a COVID-19 diagnosis (**Table 1, Supplementary Figure 2**). The median (interquartile range (IQR)) age was 49 (34-64) years. The cohort was 50.1% female and 79.6% White, 6.4% South Asian, and 2.2% Black ethnicities were recorded. Among 14,948,727 individuals in the vaccinated cohort, 858,903 (5.75%) were diagnosed with COVID-19, while of the 3,479,043 individuals in the unvaccinated cohort 149,649 (4.31%) were diagnosed with COVID-19. Individuals in the vaccinated cohort were more likely to be older, female, of white ethnicity, non-smokers, and from less deprived areas than those in unvaccinated cohort (**Table 1**). [24] Overall incidence

**Table 1:**
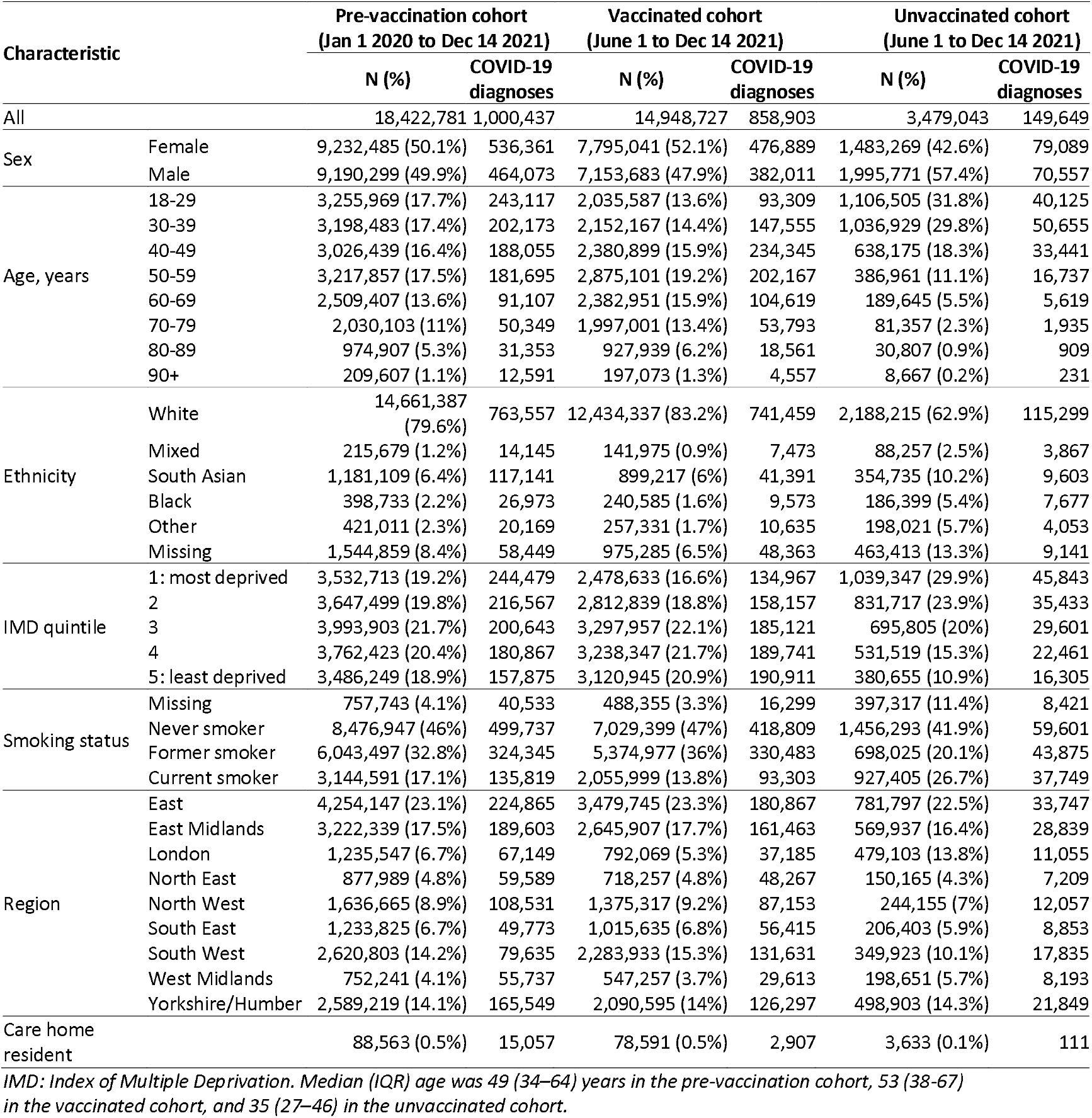
Patient characteristics in the pre-vaccination, vaccinated and unvaccinated cohorts.

| Characteristic |  | Pre-vaccination cohort<br>(Jan 1 2020 to Dec 14 2021) |  | Vaccinated cohort<br>(June 1 to Dec 14 2021) |  | Unvaccinated cohort<br>(June 1 to Dec 14 2021) |  |
| --- | --- | --- | --- | --- | --- | --- | --- |
|  |  | N (%) | COVID-19<br>diagnoses | N (%) | COVID-19<br>diagnoses | N (%) | COVID-19<br>diagnoses |
| All |  | 18,422,781 | 1,000,437 | 14,948,727 | 858,903 | 3,479,043 | 149,649 |
| Sex | Female | 9,232,485 (50.1%) | 536,361 | 7,795,041 (52.1%) | 476,889 | 1,483,269 (42.6%) | 79,089 |
|  | Male | 9,190,299 (49.9%) | 464,073 | 7,153,683 (47.9%) | 382,011 | 1,995,771 (57.4%) | 70,557 |
| Age, years | 18-29 | 3,255,969 (17.7%) | 243,117 | 2,035,587 (13.6%) | 93,309 | 1,106,505 (31.8%) | 40,125 |
|  | 30-39 | 3,198,483 (17.4%) | 202,173 | 2,152,167 (14.4%) | 147,555 | 1,036,929 (29.8%) | 50,655 |
|  | 40-49 | 3,026,439 (16.4%) | 188,055 | 2,380,899 (15.9%) | 234,345 | 638,175 (18.3%) | 33,441 |
|  | 50-59 | 3,217,857 (17.5%) | 181,695 | 2,875,101 (19.2%) | 202,167 | 386,961 (11.1%) | 16,737 |
|  | 60-69 | 2,509,407 (13.6%) | 91,107 | 2,382,951 (15.9%) | 104,619 | 189,645 (5.5%) | 5,619 |
|  | 70-79 | 2,030,103 (11%) | 50,349 | 1,997,001 (13.4%) | 53,793 | 81,357 (2.3%) | 1,935 |
|  | 80-89 | 974,907 (5.3%) | 31,353 | 927,939 (6.2%) | 18,561 | 30,807 (0.9%) | 909 |
|  | 90+ | 209,607 (1.1%) | 12,591 | 197,073 (1.3%) | 4,557 | 8,667 (0.2%) | 231 |
| Ethnicity | White | 14,661,387 (79.6%) | 763,557 | 12,434,337 (83.2%) | 741,459 | 2,188,215 (62.9%) | 115,299 |
|  | Mixed | 215,679 (1.2%) | 14,145 | 141,975 (0.9%) | 7,473 | 88,257 (2.5%) | 3,867 |
|  | South Asian | 1,181,109 (6.4%) | 117,141 | 899,217 (6%) | 41,391 | 354,735 (10.2%) | 9,603 |
|  | Black | 398,733 (2.2%) | 26,973 | 240,585 (1.6%) | 9,573 | 186,399 (5.4%) | 7,677 |
|  | Other | 421,011 (2.3%) | 20,169 | 257,331 (1.7%) | 10,635 | 198,021 (5.7%) | 4,053 |
|  | Missing | 1,544,859 (8.4%) | 58,449 | 975,285 (6.5%) | 48,363 | 463,413 (13.3%) | 9,141 |
| IMD quintile | 1: most deprived | 3,532,713 (19.2%) | 244,479 | 2,478,633 (16.6%) | 134,967 | 1,039,347 (29.9%) | 45,843 |
|  | 2 | 3,647,499 (19.8%) | 216,567 | 2,812,839 (18.8%) | 158,157 | 831,717 (23.9%) | 35,433 |
|  | 3 | 3,993,903 (21.7%) | 200,643 | 3,297,957 (22.1%) | 185,121 | 695,805 (20%) | 29,601 |
|  | 4 | 3,762,423 (20.4%) | 180,867 | 3,238,347 (21.7%) | 189,741 | 531,519 (15.3%) | 22,461 |
|  | 5: least deprived | 3,486,249 (18.9%) | 157,875 | 3,120,945 (20.9%) | 190,911 | 380,655 (10.9%) | 16,305 |
| Smoking status | Missing | 757,743 (4.1%) | 40,533 | 488,355 (3.3%) | 16,299 | 397,317 (11.4%) | 8,421 |
|  | Never smoker | 8,476,947 (46%) | 499,737 | 7,029,399 (47%) | 418,809 | 1,456,293 (41.9%) | 59,601 |
|  | Former smoker | 6,043,497 (32.8%) | 324,345 | 5,374,977 (36%) | 330,483 | 698,025 (20.1%) | 43,875 |
|  | Current smoker | 3,144,591 (17.1%) | 135,819 | 2,055,999 (13.8%) | 93,303 | 927,405 (26.7%) | 37,749 |
| Region | East | 4,254,147 (23.1%) | 224,865 | 3,479,745 (23.3%) | 180,867 | 781,797 (22.5%) | 33,747 |
|  | East Midlands | 3,222,339 (17.5%) | 189,603 | 2,645,907 (17.7%) | 161,463 | 569,937 (16.4%) | 28,839 |
|  | London | 1,235,547 (6.7%) | 67,149 | 792,069 (5.3%) | 37,185 | 479,103 (13.8%) | 11,055 |
|  | North East | 877,989 (4.8%) | 59,589 | 718,257 (4.8%) | 48,267 | 150,165 (4.3%) | 7,209 |
|  | North West | 1,636,665 (8.9%) | 108,531 | 1,375,317 (9.2%) | 87,153 | 244,155 (7%) | 12,057 |
|  | South East | 1,233,825 (6.7%) | 49,773 | 1,015,635 (6.8%) | 56,415 | 206,403 (5.9%) | 8,853 |
|  | South West | 2,620,803 (14.2%) | 79,635 | 2,283,933 (15.3%) | 131,631 | 349,923 (10.1%) | 17,835 |
|  | West Midlands | 752,241 (4.1%) | 55,737 | 547,257 (3.7%) | 29,613 | 198,651 (5.7%) | 8,193 |
|  | Yorkshire/Humber | 2,589,219 (14.1%) | 165,549 | 2,090,595 (14%) | 126,297 | 498,903 (14.3%) | 21,849 |
| Care home resident |  | 88,563 (0.5%) | 15,057 | 78,591 (0.5%) | 2,907 | 3,633 (0.1%) | 111 |

IMD: Index of Multiple Deprivation. Median (IQR) age was 49 (34–64) years in the pre-vaccination cohort, 53 (38–67) in the vaccinated cohort, and 35 (27–46) in the unvaccinated cohort.

In each cohort, the incidence of gastrointestinal disease was higher after a diagnosis of COVID-19 than before or without a COVID-19 diagnosis, with the highest incidence rates after hospitalised COVID-19 (**Table 2**). In the pre-vaccination cohort, the total number of events (total after COVID-19) were: upper gastrointestinal bleeding 143,769 (7,764); lower gastrointestinal bleeding 177,909 (8,526); acute pancreatitis 26,937 (1,314); appendicitis 27,801 (1,290); gastro-oesophageal reflux 413,727 (19,680); dyspepsia 165,399 (7,254); peptic ulcer 24,957 (1,122); nonalcoholic steatohepatitis 4,443 (300); gallstones 152,367 (7,164); and irritable bowel syndrome 151,527 (7,314).

**Table 2.**
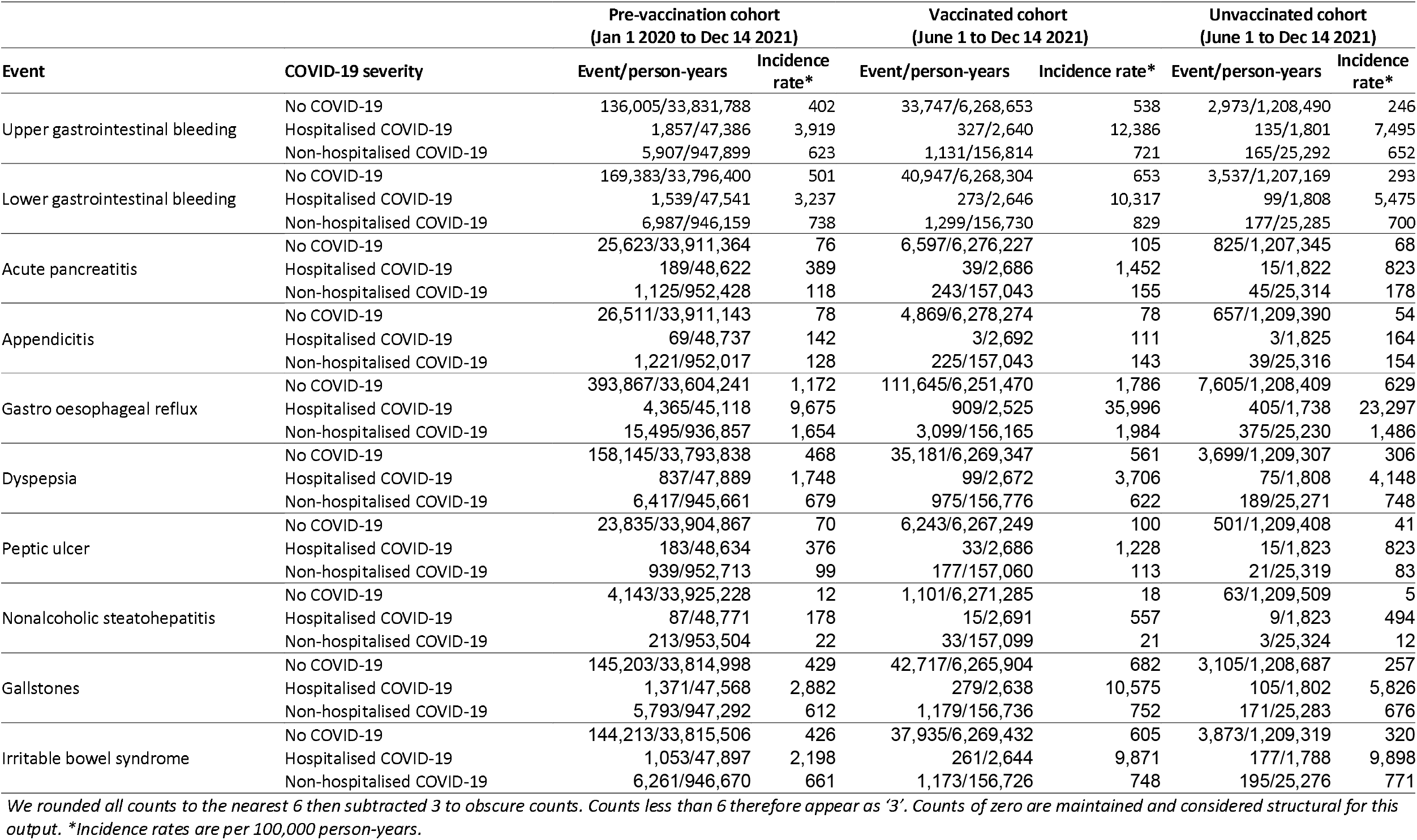
Number of gastrointestinal disease events in pre-vaccination, vaccinated and unvaccinated cohorts, with person-years of follow-up, by COVID-19 severity.

### Comparisons of event rates after COVID-19 diagnosis versus before or without COVID-19

Maximally adjusted hazard ratios (aHRs) comparing the incidence of each outcome post– COVID-19 with incidence before or without COVID-19 did not differ substantially from age- and sex–adjusted HRs in any cohort (**Table 3; Supplementary Table 3**). Incidence of all outcomes was extremely high on day 0 (Table 3).

**Table 3:** Adjusted hazard ratios (95% CI) comparing the incidence of gastrointestinal disease events after COVID-19 with the incidence before or without COVID-19, in the pre-vaccination, vaccinated and unvaccinated cohorts. Hazard ratios are maximally adjusted.

| Outcome | Time since COVID-19 | Pre-vaccination cohort | Vaccinated cohort | Unvaccinated cohort |
| --- | --- | --- | --- | --- |
| Upper gastrointestinal bleeding | Day 0 | 177 (168-186) | 52.7 (48.3-57.6) | 106 (87.6-128) |
|  | Weeks 1-4 | 3.77 (3.53-4.03) | 1.84 (1.67-2.03) | 4.26 (3.51-5.19) |
|  | Weeks 5-28 | 1.59 (1.52-1.65) | 1.19 (1.10-1.30) | 1.40 (1.10-1.77) |
|  | Weeks 29-52 | 1.29 (1.23-1.35) | - | - |
|  | Weeks 53-102 | 1.43 (1.33-1.54) | - | - |
| Lower gastrointestinal bleeding | Day 0 | 91.6 (86.5-97.1) | 29.3 (26.5-32.4) | 55.5 (44.5-69.2) |
|  | Weeks 1-4 | 2.47 (2.31-2.65) | 1.41 (1.28-1.55) | 2.85 (2.32-3.50) |
|  | Weeks 5-28 | 1.46 (1.41-1.52) | 1.19 (1.11-1.28) | 1.34 (1.09-1.65) |
|  | Weeks 29-52 | 1.25 (1.20-1.30) | - | - |
|  | Weeks 53-102 | 1.35 (1.26-1.45) | - | - |
| Acute pancreatitis | Day 0 | 109 (94.7-126) | 42.1 (34.1-52.1) | 50.0 (31.4-79.4) |
|  | Weeks 1-4 | 2.80 (2.38-3.30) | 1.70 (1.37-2.11) | 2.75 (1.82-4.16) |
|  | Weeks 5-28 | 1.33 (1.21-1.47) | 1.12 (0.92-1.35) | 1.14 (0.72-1.80) |
|  | Weeks 29-52 | 1.22 (1.10-1.36) | - | - |
|  | Weeks 53-102 | 1.63 (1.39-1.92) | - | - |
| Appendicitis | Day 0 | 50.5 (42.2-60.4) | 17.7 (12.8-24.5) | 31.0 (16.1-59.5) |
|  | Weeks 1-4 | 1.79 (1.49-2.16) | 1.59 (1.26-2.00) | 2.43 (1.48-4.01) |
|  | Weeks 5-28 | 1.23 (1.13-1.34) | 1.30 (1.08-1.57) | 1.78 (1.13-2.81) |
|  | Weeks 29-52 | 1.18 (1.07-1.30) | - | - |
|  | Weeks 53-102 | 1.24 (1.02-1.50) | - | - |
| Gastro oesophageal reflux | Day 0 | 70.8 (67.8-74.0) | 18.6 (17.3-20.1) | 55.9 (48.5-64.5) |
|  | Weeks 1-4 | 3.36 (3.23-3.50) | 1.42 (1.34-1.51) | 3.81 (3.39-4.29) |
|  | Weeks 5-28 | 1.35 (1.32-1.38) | 1.20 (1.15-1.25) | 1.65 (1.46-1.87) |
|  | Weeks 29-52 | 1.21 (1.18-1.24) | - | - |
|  | Weeks 53-102 | 1.33 (1.28-1.39) | - | - |
| Dyspepsia | Day 0 | 28.4 (25.7-31.4) | 7.31 (5.98-8.94) | 31.3 (24.2-40.5) |
|  | Weeks 1-4 | 1.65 (1.52-1.79) | 1.04 (0.93-1.16) | 2.21 (1.79-2.73) |
|  | Weeks 5-28 | 1.25 (1.21-1.30) | 1.13 (1.04-1.22) | 1.53 (1.27-1.84) |
|  | Weeks 29-52 | 1.22 (1.17-1.27) | - | - |
|  | Weeks 53-102 | 1.25 (1.16-1.34) | - | - |
| Peptic ulcer | Day 0 | 50.4 (40.4-63.0) | 15.9 (11.2-22.6) | 33.2 (14.8-74.8) |
|  | Weeks 1-4 | 4.38 (3.75-5.12) | 2.20 (1.78-2.73) | 2.97 (1.66-5.31) |
|  | Weeks 5-28 | 1.73 (1.57-1.90) | 1.20 (0.99-1.47) | 1.97 (1.18-3.31) |
|  | Weeks 29-52 | 1.30 (1.16-1.45) | - | - |
|  | Weeks 53-102 | 1.30 (1.08-1.57) | - | - |
| Non-alcoholic steatohepatitis | Day 0 | 258 (198-337) | 26.4 (14.5-48.0) | X |
|  | Weeks 1-4 | 5.89 (4.01-8.66) | 2.17 (1.40-3.36) | X |
|  | Weeks 5-28 | 1.57 (1.05-2.35) | 0.93 (0.59-1.48) | X |
|  | Weeks 29-52 | 1.10 (0.73-1.65) | - | - |
|  | Weeks 53-102 | 1.75 (0.95-3.22) | - | - |
| Gallstones | Day 0 | 98.9 (92.8-105) | 26.3 (23.6-29.2) | 41.9 (32.5-54.2) |
|  | Weeks 1-4 | 2.73 (2.53-2.94) | 1.38 (1.25-1.52) | 2.89 (2.34-3.57) |
|  | Weeks 5-28 | 1.45 (1.39-1.51) | 1.10 (1.01-1.18) | 1.72 (1.41-2.09) |
|  | Weeks 29-52 | 1.21 (1.16-1.26) | - | - |
|  | Weeks 53-102 | 1.35 (1.25-1.45) | - | - |
| Irritable bowel syndrome | Day 0 | 47.5 (44.0-51.3) | 13.7 (12.1-15.5) | 37.1 (29.8-46.2) |
|  | Weeks 1-4 | 2.20 (2.04-2.37) | 1.17 (1.06-1.29) | 3.39 (2.90-3.96) |
|  | Weeks 5-28 | 1.14 (1.10-1.19) | 1.09 (1.01-1.16) | 1.38 (1.15-1.65) |
|  | Weeks 29-52 | 1.12 (1.08-1.17) | - | - |
|  | Weeks 53-102 | 1.15 (1.07-1.23) | - | - |

*X represents insufficient events for estimation*

### Upper gastrointestinal bleeding

Incidence of upper gastrointestinal bleeding was elevated during weeks 1–4 post–COVID-19, compared with before or without COVID-19: aHRs were 3.77 (95% CI 3.53–4.03) in the pre-vaccination cohort, 1.84 (1.67–2.03) in the vaccinated cohort, and 4.26 (3.51–5.19) in the unvaccinated cohort (**Figure 1; Table 3**). In the pre-vaccination cohort, incidence remained elevated through weeks 53–102: aHR 1.43 (1.33–1.54). During weeks 1–4, aHRs were substantially higher after hospitalisation with COVID-19 (pre-vaccination: 20.1 (18.4–22.1); vaccinated: 15.2 (13.4–17.3); unvaccinated: 19.7 (15.0–25.9)) than after non-hospitalised COVID-19 (pre-vaccination: 1.92 (1.74–2.11); vaccinated: 1.20 (1.06–1.36); unvaccinated: 2.14 (1.61–2.85)) (**Figure 1; Supplementary Tables 4 and 5**). In the pre-vaccination cohort, aHRs remained higher after hospitalised than non-hospitalised COVID-19, throughout follow-up.

**Figure1:**
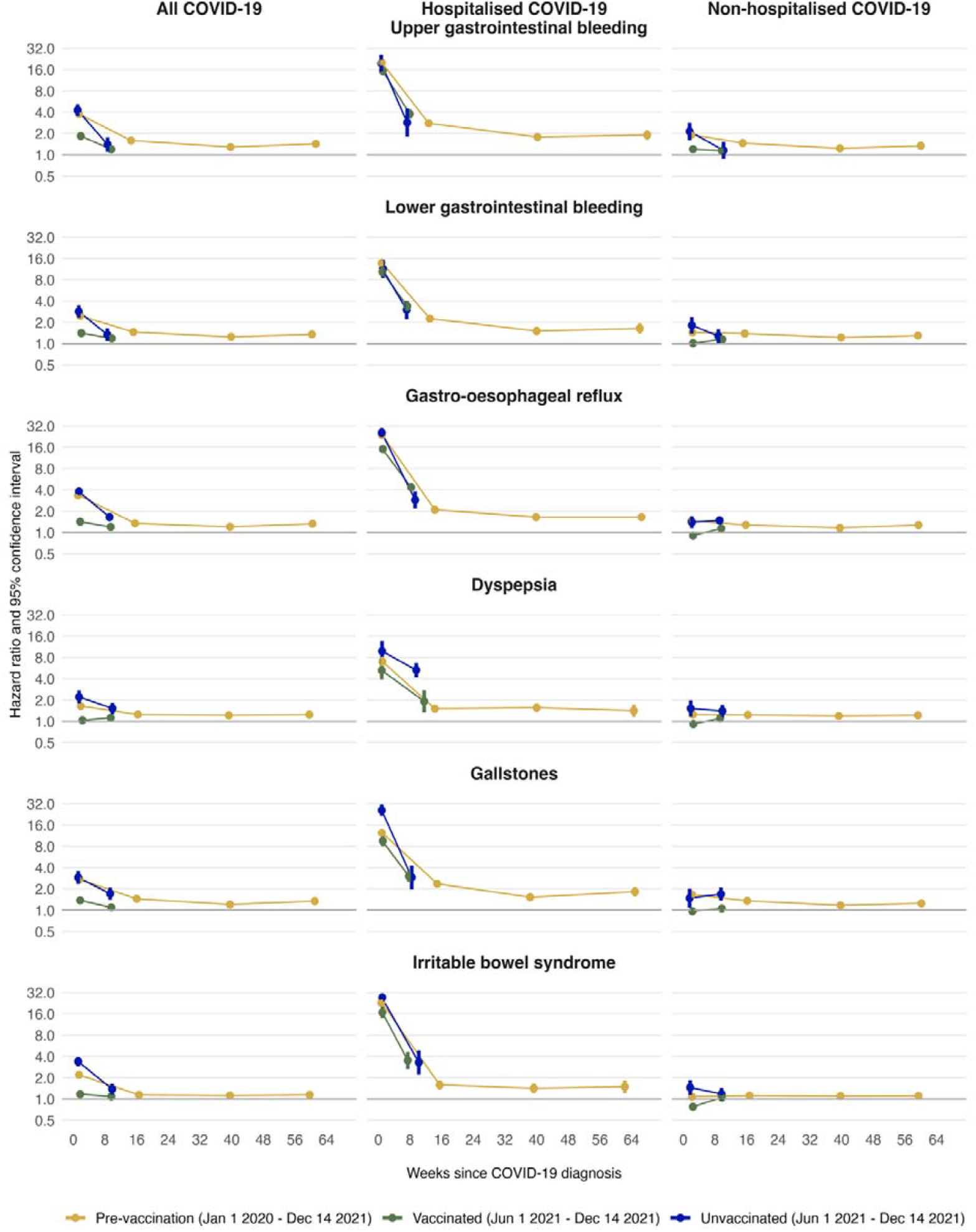
Maximally adjusted hazard ratios and 95% CIs comparing the incidence of gastrointestinal disease events after COVID-19 with the incidence before or without COVID-19, in the pre-vaccination, vaccinated and unvaccinated cohorts, overall and by COVID-19 severity with day 0 excluded.

### Lower gastrointestinal bleeding

Across all cohorts, the incidence of lower gastrointestinal bleeding was elevated during weeks 1–4 post–COVID-19 compared with before or without COVID-19: pre-vaccination: 2.47 (95% CI 2.31–2.65); vaccinated: 1.41 (1.28–1.55); unvaccinated: 2.85 (2.32–3.50) (**Figure 1; Table 3**). In the pre-vaccination cohort, incidence remained elevated through weeks 53–102 (aHR 1.35 (1.26–1.45)). During weeks 1–4, aHRs were substantially higher after hospitalised COVID-19 (pre-vaccination: 13.7 (12.4–15.2); vaccinated: 10.4 (8.80– 12.2); unvaccinated: 11.4 (8.48–15.4)) than after non-hospitalised COVID-19 (pre-vaccination: 1.44 (1.32–1.59); vaccinated: 1.02 (0.91–1.14); unvaccinated: 1.82 (1.39– 2.38)) (**Figure 1; Supplementary Tables 4 and 5**). In the pre-vaccination cohort, aHRs remained higher after hospitalised than non-hospitalised COVID-19, throughout follow-up.

### Subgroup analysis

Subgroup analyses showed notable variation in aHRs post–COVID-19 by age and by prior gastrointestinal events, whereas differences by sex, ethnicity, prior gastrointestinal events and prior gastrointestinal operations were minimal (**Supplementary tables 4-17, Supplementary Figures 3-7**). Adjusted HRs were consistently higher in older versus younger age groups (**Supplementary tables 6-9, Supplementary Figure 3**). For example, in the pre-vaccination cohort during weeks 1–4, aHRs for upper gastrointestinal bleeding rose from 1.69 (95% CI 1.40–2.04) in individuals aged 18–39 to 5.04 (4.37–5.81) in those aged 60–79. A similar pattern was seen for lower gastrointestinal bleeding, with aHRs increasing from 1.16 (0.99–1.37) to 3.96 (3.49–4.49) across the same age groups. These age-related differences persisted through weeks 5–28 post-diagnosis.

Adjusted HRs were higher for individuals without than with prior gastrointestinal events (**Supplementary tables 14-15, Supplementary Figure 6**). In the pre-vaccination cohort, for upper gastrointestinal bleeding during weeks 1–4, the aHRs for upper gastrointestinal bleeding during weeks 1-4 were 5.36 (95% CI 4.78-6.01) and 3.36 (3.09-3.64) among those without and with prior events respectively. Similar patterns were observed for lower gastrointestinal bleeding (without prior event: 3.00 (2.65-3.40); with prior event: 2.31 (2.13-2.51)).

### Other gastrointestinal diseases

aHRs for other gastrointestinal diseases were broadly similar to those for upper and lower gastrointestinal bleeding, both overall (**Table 3**) and by COVID-19 severity (**Figure 3; Supplementary Tables 4 and 5**). As with upper and lower gastrointestinal bleeding, aHRs for these diseases were attenuated in the vaccinated compared with the pre-vaccination and unvaccinated cohorts during weeks 1–4 post–COVID-19.

### Absolute excess risk

Estimated excess risks of upper gastrointestinal bleeding 28 weeks after COVID-19, standardised to the age distribution of the pre-vaccination cohort, were 159 per 100,000 people diagnosed with COVID-19 in the pre-vaccination cohort, 60 in the vaccinated cohort, and 112 in the unvaccinated cohort (**Supplementary Table 18, Supplementary Figure 8**). For lower gastrointestinal bleeding, estimated excess risks 28 weeks post-COVID-19 were 144 per 100,000 people diagnosed with COVID-19 in the pre-vaccination cohort, 64 in the vaccinated cohort, and 91 in the unvaccinated cohort. (**Supplementary Table 18, Supplementary Figure 9**). In all cohorts, for upper and lower gastrointestinal bleeding, excess risks increased with age: the oldest group (80–110 years) consistently had the highest risks, while the youngest group (18–39 years) had the lowest. Similar patterns were observed for other gastrointestinal outcomes (**Supplementary Figures 10–13**).

## DISCUSSION

### Main findings

In this population-based study of over 18 million people followed for up to two years, COVID-19 was associated with marked elevations in the incidence of gastrointestinal diseases during the first month after diagnosis in the pre-vaccination and unvaccinated cohorts. While associations declined with increasing time since COVID-19 diagnosis, the incidence of gastrointestinal disease remained higher for up to 2 years post-COVID-19 diagnosis in the pre-vaccination cohort and for 6-months in the unvaccinated cohort.

Incidence of gastrointestinal diseases after COVID-19 was elevated in the vaccinated cohort, but to a lesser extent than in the other cohorts. Overall differences between the vaccinated and unvaccinated cohorts were largely explained by whether COVID-19 led to hospitalisation, with the elevation in incidence of gastrointestinal diseases much greater after hospitalised than non-hospitalised COVID-19 in all cohorts. Amongst vaccinated individuals, there was little elevation in the incidence of gastrointestinal diseases after non-hospitalised COVID-19. Patterns of association were similar across all gastrointestinal diseases studied.

### Comparison with literature

Our findings are consistent with previous studies reporting an association of COVID-19 diagnosis with higher rates of long-term gastrointestinal diseases, including gastroesophageal reflux, dyspepsia, IBS, gastrointestinal bleeding and acute pancreatitis. [12,20,25–27] We also observed associations with gallstones, peptic ulcer, and non-alcoholic steatohepatitis, consistent with results reported in recent studies and meta-analyses. [4,13,19,25] Another study reported gastrointestinal manifestations in the acute phase of infection across multiple cohorts. [13] Additionally, systematic reviews have reported gastrointestinal bleeding following COVID-19, particularly among hospitalised individuals receiving anticoagulants, with prevalence ranging between 0.5% and 3% depending on disease severity. [25,26]

Our findings are consistent with those of Xu et al.[18], who identified an increased risk of a broad range of gastrointestinal diseases post-COVID-19, particularly among those with severe illness, using US Veterans Affairs data. However, that cohort was composed mostly of older unvaccinated men, limiting generalisability. Our study used a large, nationally representative dataset from England, and accounted for vaccination status, COVID-19 severity and different population subgroups.

The attenuation of post-COVID gastrointestinal rates among vaccinated individuals is likely due to a reduction of disease severity following immunisation, consistent with studies showing lower risk of long COVID among vaccinated people. [22,28] Breakthrough infection studies confirm that vaccinated individuals can still develop gastrointestinal symptoms, although the risk remains lower compared to unvaccinated individuals. [29] This indicates that, while vaccination is unlikely to fully prevent post-COVID gastrointestinal complications, it may substantially reduce the risk of these outcomes.

### Clinical implications

Findings from this study are consistent with COVID-19 being a systemic illness that affects multiple systems. Severely affected patients are more likely to have gastrointestinal acute diseases, including upper and lower gastrointestinal bleeding. Our results further highlight the importance of vaccination in reducing the risk of severe COVID-19, and thus the subsequent risk of long-term harm.

### Strengths and limitations

Strengths of this study include the large and representative study population, the availability of detailed linked electronic health record data, the relatively long duration of follow-up, and the opportunity to examine the role of vaccination in the relationship between COVID-19 and gastrointestinal diseases. We also note several limitations. First, those who were unvaccinated may have been less likely to contact health services and to test for SARS-CoV-2, which might have led to underestimated effects in unvaccinated people not hospitalised with COVID-19. Those with recorded COVID-19, particularly those hospitalised, may have been more likely to have their gastrointestinal disease recorded due to greater contact with health services. This may have underpinned the particularly high HRs observed initially following diagnosis, especially in those hospitalised, and the rapid fall thereafter as service contact is likely to be highest in the early post-diagnosis period. However, such issues are unlikely to fully explain the adverse effect of COVID-19 on gastrointestinal diseases, given the persistent elevation in incident outcomes after hospitalised COVID-19 and the variation in associations with different diseases. Conversely, under ascertainment of gastrointestinal diseases due to reduced use of primary care services during the pandemic and cessation of normal screening activities, may have biased associations towards the null. A further limitation is that we could only assess COVID-19 severity according to whether patients were hospitalised and did not consider the potential role of repeated infections. The effect of different COVID-19 variants on subsequent gastrointestinal diagnosis could not be directly accounted for as individual-level information on variant was not available. However, the cohorts were aligned to periods when specific variants were dominant in the population. We observed similar patterns in the pre-vaccinated and unvaccinated cohort, when different COVID-19 variants were dominant and treatment strategies had evolved, indicating that variants may have a limited impact on gastrointestinal outcomes and findings could extend to more recent infection waves. Finally, we cannot exclude the possibility of unmeasured confounding, although we controlled for a wide range of demographic characteristics and prior morbidities.

## Conclusions

The incidence of gastrointestinal diseases is elevated for up to two years after diagnosis of COVID-19, particularly severe COVID-19 leading to hospitalisation. Our findings support the role of vaccination against COVID-19 in mitigating the long-term effect of COVID-19 on gastrointestinal health.

## Data Availability

All analytic code, codelists, and study definitions are openly available at: https://github.com/opensafely/post-covid-gastrointestinal
. Aggregate, non-disclosive outputs are provided in the manuscript and Supplementary Materials. The underlying patient-level data are pseudonymised and held within the NHS England OpenSAFELY platform; they cannot be shared publicly for legal and privacy reasons. Accredited researchers may apply for controlled access via OpenSAFELY under NHS England approvals; see https://www.opensafely.org
for access details.

## FUNDING

This study was supported by the COVID-19 Longitudinal Health and Wellbeing National Core Study, funded by the UKRI Medical Research Council (MC_PC_20059); the COVID-19 Data and Connectivity National Core Study, funded by the UKRI Medical Research Council; and the CONVALESCENCE long COVID study, funded by the UK National Institute for Health and Care Research (COVID-LT-009). MA is supported by Phase 1 Covid-19 Longitudinal Health and Wellbeing – National Core Study funded by the Medical Research Council (MC_PC_20059); the CONVALESCENCE programme funded by NIHR (COV-LT-0009); the HDR UK Medicines Driver Programme. GC is supported by Health Data Research UK, the British Heart Foundation, the BHF Cambridge Centre of Research Excellence and the NIHR Cambridge Biomedical Research Centre. RD is supported by the NIHR Bristol Biomedical Research Centre, NIHR (“Impact and inequalities of winter pressures in primary care”), Health Data Research UK South West and the NHS Data for Research & Development South West Secure Data Environment. VW is supported by the MRC Integrative Epidemiology Unit at the University of Bristol (MC_UU_00032/03), the EU Horizon programme and NIHR, including NIHR Research for Patient Benefit and NIHR funding for work on evaluation of COVID-19 vaccine products and schedules and winter pressures in primary care. NC receives research funding from NIHR and UKRI paid to her institution. SB is supported by the Bennett Foundation, NHS England, the NIHR Oxford Biomedical Research Centre, the Wellcome Trust, XTX Markets and Health Data Research UK. YW was supported by a UKRI MRC fellowship (MC/W021358/1).

The OpenSAFELY platform is principally funded by grants from:

- NHS England [2023-2025];
- The Wellcome Trust (222097/Z/20/Z) [2020-2024];
- MRC (MR/V015737/1) [2020-2021].

Additional contributions to OpenSAFELY have been funded by grants from:

- MRC via the National Core Study programme, Longitudinal Health and Wellbeing strand (MC_PC_20030, MC_PC_20059) [2020-2022] and the Data and Connectivity strand (MC_PC_20058) [2021-2022];
- NIHR and MRC via the CONVALESCENCE programme (COV-LT-0009, MC_PC_20051) [2021-2024];
- NHS England via the Primary Care Medicines Analytics Unit [2021-2024].

## INFORMATION GOVERNANCE AND ETHICAL APPROVAL

NHS England is the data controller of the NHS England OpenSAFELY COVID-19 Service; TPP is the data processor; all study authors using OpenSAFELY have the approval of NHS England.[30] This implementation of OpenSAFELY is hosted within the TPP environment which is accredited to the ISO 27001 information security standard and is NHS IG Toolkit compliant;[31]

Patient data has been pseudonymised for analysis and linkage using industry standard cryptographic hashing techniques; all pseudonymised datasets transmitted for linkage onto OpenSAFELY are encrypted; access to the NHS England OpenSAFELY COVID-19 service is via a virtual private network (VPN) connection; the researchers hold contracts with NHS England and only access the platform to initiate database queries and statistical models; all database activity is logged; only aggregate statistical outputs leave the platform environment following best practice for anonymisation of results such as statistical disclosure control for low cell counts.[32]

The service adheres to the obligations of the UK General Data Protection Regulation (UK GDPR) and the Data Protection Act 2018. The service previously operated under notices initially issued in February 2020 by the Secretary of State under Regulation 3(4) of the Health Service (Control of Patient Information) Regulations 2002 (COPI Regulations), which required organisations to process confidential patient information for COVID-19 purposes; this set aside the requirement for patient consent.[33] As of 1 July 2023, the Secretary of State has requested that NHS England continue to operate the Service under the COVID-19 Directions 2020.[34] In some cases of data sharing, the common law duty of confidence is met using, for example, patient consent or support from the Health Research Authority Confidentiality Advisory Group.[35]

Taken together, these provide the legal bases to link patient datasets using the service. GP practices, which provide access to the primary care data, are required to share relevant health information to support the public health response to the pandemic, and have been informed of how the service operates.

This study was approved by the Health Research Authority [REC reference 22/PR/0095] and by the University of Bristol’s Faculty of Health Sciences Ethics Committee [reference 117269]

## PATIENT AND PUBLIC INVOLVEMENT AND ENGAGEMENT (PPIE)

OpenSAFELY has involved patients and the public in various ways: we developed a public website that provides a detailed description of the platform in language suitable for a lay audience (https://opensafely.org); we have participated in two citizen juries exploring public trust in OpenSAFELY; we have co-developed an explainer video (https://www.opensafely.org/about/); we have patient representation who are experts by experience on our OpenSAFELY Oversight Board; we have partnered with Understanding Patient Data to produce lay explainers on the importance of large datasets for research; we have presented at various online public engagement events to key communities (e.g., Healthcare Excellence Through Technology; Faculty of Clinical Informatics annual conference; NHS Assembly; HDRUK symposium); and more. To ensure the patient voice is represented, we are working closely to decide on language choices with appropriate medical research charities (e.g., Association of Medical Research Charities). We will share information and interpretation of our findings through press releases, social media channels, and plain language summaries.

## ACKNOWLEDGEMENT CONTENT

We are very grateful for all the support received from the TPP Technical Operations team throughout this work, and for generous assistance from the information governance and database teams at NHS England and the NHS England Transformation Directorate.

