## Supplementary materials for "The impact of COVID-19 on gastrointestinal diseases in vaccinated and unvaccinated individuals: a population-based cohort study in England"

### Supplementary Methods

#### Absolute excess risk

The absolute excess risk analysis was performed for each outcome in each cohort using the hazard ratio from the main analysis. To compare the outcomes across the cohorts, each of which have different lengths of follow-up, we calculated the absolute excess risk at 28 weeks.  We accounted for age in the analysis by using event counts and person-days from four age groups: 18–39, 40–59, 60–79, and 80–110 years. For each group, we calculated the average daily incidence of the outcome in the unexposed and the cumulative risk over time. We then used the relevant hazard ratio for that day (e.g., days 0 to 27 will have the coefficient for the term ‘days0_28’, while days 28 to 196 will have the coefficient for the term ‘days28_197’) to predict the expected cumulative survival in the exposed. Finally, we calculated the daily excess risk as the difference in cumulative survival for the unexposed and the expected cumulative survival in the exposed. The overall absolute excess risk was estimated using a weighted sum of the excess risks in each group, weighted by the proportions of individuals in age strata in the pre-vaccine availability cohort.

#### Information governance

NHS England is the data controller of the NHS England OpenSAFELY COVID-19 Service; TPP is the data processor; all study authors using OpenSAFELY have the approval of NHS England. This implementation of OpenSAFELY is hosted within the TPP environment which is accredited to the ISO 27001 information security standard and is NHS IG Toolkit compliant.

Patient data have been pseudonymised for analysis and linkage using industry standard cryptographic hashing techniques; all pseudonymised datasets transmitted for linkage onto OpenSAFELY are encrypted; access to the NHS England OpenSAFELY COVID-19 service is via a virtual private network (VPN) connection; the researchers hold contracts with NHS England and only access the platform to initiate database queries and statistical models; all database activity is logged; only aggregate statistical outputs leave the platform environment following best practice for anonymisation of results such as statistical disclosure control for low cell counts.

The service adheres to the obligations of the UK General Data Protection Regulation (UK GDPR) and the Data Protection Act 2018. The service previously operated under notices initially issued in February 2020 by the Secretary of State under Regulation 3(4) of the Health Service (Control of Patient Information) Regulations 2002 (COPI Regulations), which required organisations to process confidential patient information for COVID-19 purposes; this set aside the requirement for patient consent. As of 1 July 2023, the Secretary of State has requested that NHS England continue to operate the Service under the COVID-19 Directions 2020. In some cases of data sharing, the common law duty of confidence is met using, for example, patient consent or support from the Health Research Authority Confidentiality Advisory Group.

Taken together, these provide the legal bases to link patient datasets using the service. GP practices, which provide access to the primary care data, are required to share relevant health information to support the public health response to the pandemic and have been informed of how the service operates.

This study was approved by the Health Research Authority [REC reference 22/PR/0095] and by the University of Bristol's Faculty of Health Sciences Ethics Committee [reference 117269].

### Supplementary Tables

#### Supplementary Table 1. Summary of covariate definitions

| Confounder | Type | Definition |
| --- | --- | --- |
| Sex | Categorical | Male, Female |
| Age | Continuous | Modelled as age in years using a restricted cubic spline with 3 knots at the 10^th^, 50^th^ and 90^th^ percentiles |
| Ethnicity | Categorical | 1: White  2: Mixed  3: South Asian  4: Black  5: Other  Missing |
| Deprivation | Categorical | 10 categories from Index of Multiple Deprivation 2019 |
| Region | Categorical | East of England  London  Midlands  North East and Yorkshire  North West  South East  South West |
| Consultation rate | Continuous | Number of primary care contacts in the year prior to pandemic start (2019). |
| Smoking status | Categorical | E: Ever smoker  M: Missing  N: Never smoker  S: Current smoker |
| Obesity | Binary | 1: if BMI>=30 or coded diagnosis for obesity; 0: otherwise |
| Healthcare worker | Binary | 1: if healthcare worker; 0: otherwise |
| Care home resident | Binary | 1: if care home resident; 0: otherwise |
| Alcohol above limits | Binary | 1: exceeds limits; 0: within limits |
| Cholelithiasis | Binary | 1: diagnosis present; 0: otherwise |
| H pylori infection | Binary | 1: diagnosis present; 0: otherwise |
| NSAIDS medication | Binary | 1: if prescription present; 0: otherwise |
| Aspirin medication | Binary | 1: if prescription present; 0: otherwise |
| Anticoagulants medications | Binary | 1: if prescription present; 0: otherwise |
| Antidepressants | Binary | 1: if prescription present; 0: otherwise |
| Prior symptoms and gastrointestinal diseases | Binary | 1: if diagnosis; 0: otherwise |
| Previous gastrointestinal operation | Binary | 1: if existing operation; 0: otherwise |
| Hypercalcemia | Binary | 1: diagnosis present, 0: otherwise |
| Hypertriglyceridemia | Binary | 1: diagnosis present, 0: otherwise |
| HDL level | Continuous |  |
| Systolic blood pressure | Continuous |  |

#### Supplementary Table 2: Summary of cohort characteristics.

| Characteristic | Pre-vaccination cohort | Vaccinated cohort | Unvaccinated cohort |
| --- | --- | --- | --- |
| Start date | 01/01/2020, which is the approximate start date of the pandemic in the UK. | 01/06/2021, which is the date that the delta variant was thought to be ubiquitous in England. | 01/06/2021, which is the date that the delta variant was thought to be ubiquitous in England. |
| End date - exposure | 18/06/2021, which is the date when the Joint Committee for Vaccination and Immunisation (JCVI) phase 2, group 12 (all adults aged 18 years and older) become eligible for a COVID-19 vaccination. | 14/12/2021, which is the day that the UK Health Security Agency stated that over half of English cases they sampled have S Gene Target Failure, meaning they were likely Omicron.* | 14/12/2021, which is the day that the UK Health Security Agency stated that over half of English cases they sampled have S Gene Target Failure, meaning they were likely Omicron.* |
| End date - outcome | 14/12/2021, which is the day that the UK Health Security Agency stated that over half of English cases they sampled have S Gene Target Failure, meaning they were likely Omicron.* | 14/12/2021, which is the day that the UK Health Security Agency stated that over half of English cases they sampled have S Gene Target Failure, meaning they were likely Omicron.* | 14/12/2021, which is the day that the UK Health Security Agency stated that over half of English cases they sampled have S Gene Target Failure, meaning they were likely Omicron.* |
| Exclusion criteria | Patients will be excluded if they meet any of the following criteria:  COVID-19 diagnosis recorded prior to their index date | Patients will be excluded if they meet any of the following criteria:  COVID-19 diagnosis recorded prior to their index date [Note: these individuals are required for a sensitivity analysis and so should not be removed at the data extraction stage]  They do not have a record of two vaccination doses prior to the study end date  They received a vaccination prior to 08-12-2020 (i.e., the start of the vaccination program)  They received a second dose vaccination before their first dose vaccination  They received a second dose vaccination less than three weeks after their first dose  They received mixed vaccine products before 07-05-2021 | Patients will be excluded if they meet any of the following criteria:  COVID-19 diagnosis recorded prior to their index date [Note: these individuals are required for a sensitivity analysis and so should not be removed at the data extraction stage]  They have a record of one or more vaccination doses prior to their index date  They could not be assigned to a vaccination group as defined by the Joint Committee on Vaccination and Immunisation (JCVI) |
| Follow-up start | Study start date. | Follow-up will start at the latest of the following dates (i.e., an individual’s index date): Two weeks after their second vaccination; Study start date. | Follow-up will start at the latest of the following dates (i.e., an individual’s index date):  12 weeks after they became eligible for vaccination; Study start date. |
| Follow-up end for exposure | Follow-up will end at the earliest of the following dates: Death; Outcome event; Study end date exposure; Deregistration date; Vaccination; Date when eligible for vaccination according to JCVI priority groupings. | Follow-up will end at the earliest of the following dates: Death; Outcome event; Study end date exposure; Deregistration date. | Follow-up will end at the earliest of the following dates: Death; Outcome event; Study end date exposure; Deregistration date; Vaccination. |
| Follow-up end for outcomes | Follow-up will end at the earliest of the following dates: Death; Outcome event; Study end date outcome; Deregistration date. | Follow-up will end at the earliest of the following dates: Death; Outcome event; Study end date outcome; Deregistration date. | Follow-up will end at the earliest of the following dates: Death; Outcome event; Study end date outcome; Deregistration date. |
| Cox regression time periods | [0,28), [28,197), [197, 365), [365,714) | [0,28), [28,197) | [0,28), [28,197) |
| * UK Health Security Agency. (2021). *Omicron daily overview: 17 December 2021*. Retrieved 26 April 2024, from <https://assets.publishing.service.gov.uk/government/uploads/system/uploads/attachment_data/file/1042100/20211217_OS_Daily_Omicron_Overview.pdf> | | | |

#### Supplementary Table 3: Age-sex adjusted hazard ratios (95% CI) comparing the incidence of gastrointestinal disease events after COVID-19 with the incidence before or without COVID-19, in the pre-vaccination, vaccinated and unvaccinated cohorts.

X insufficient events

| Outcome | Time since COVID-19 | Pre-vaccination cohort | Vaccinated cohort | Unvaccinated cohort |
| --- | --- | --- | --- | --- |
| Upper gastrointestinal bleeding | Day 0 | 202 (192-212) | 55.3 (50.6-60.4) | 138 (114-166) |
|  | Weeks 1-4 | 4.44 (4.16-4.73) | 1.95 (1.77-2.15) | 5.65 (4.66-6.85) |
|  | Weeks 5-28 | 1.79 (1.72-1.86) | 1.30 (1.20-1.42) | 1.92 (1.52-2.43) |
|  | Weeks 29-52 | 1.45 (1.39-1.52) | - | - |
|  | Weeks 53-102 | 1.74 (1.62-1.87) | - | - |
| Lower gastrointestinal bleeding | Day 0 | 104 (98.3-110) | 31.3 (28.4-34.5) | 74.6 (59.9-92.9) |
|  | Weeks 1-4 | 2.77 (2.59-2.97) | 1.50 (1.36-1.65) | 3.86 (3.14-4.73) |
|  | Weeks 5-28 | 1.63 (1.58-1.69) | 1.30 (1.20-1.39) | 1.92 (1.56-2.36) |
|  | Weeks 29-52 | 1.40 (1.35-1.46) | - | - |
|  | Weeks 53-102 | 1.61 (1.50-1.72) | - | - |
| Acute pancreatitis | Day 0 | 137 (120-157) | 44.3 (35.9-54.6) | 72.0 (45.5-114) |
|  | Weeks 1-4 | 3.21 (2.71-3.81) | 1.78 (1.43-2.22) | 4.02 (2.67-6.06) |
|  | Weeks 5-28 | 1.50 (1.36-1.66) | 1.21 (1.00-1.47) | 1.82 (1.16-2.86) |
|  | Weeks 29-52 | 1.38 (1.24-1.53) | - | - |
|  | Weeks 53-102 | 1.97 (1.68-2.32) | - | - |
| Appendicitis | Day 0 | 53.3 (44.6-63.8) | 18.1 (13.0-25.0) | 37.3 (19.5-71.5) |
|  | Weeks 1-4 | 1.89 (1.57-2.28) | 1.64 (1.31-2.06) | 2.96 (1.80-4.87) |
|  | Weeks 5-28 | 1.30 (1.19-1.42) | 1.36 (1.13-1.64) | 2.25 (1.43-3.54) |
|  | Weeks 29-52 | 1.25 (1.14-1.37) | - | - |
|  | Weeks 53-102 | 1.34 (1.11-1.63) | - | - |
| Gastro-oesophageal reflux | Day 0 | 85.6 (82.1-89.3) | 19.7 (18.3-21.2) | 79.2 (68.8-91.3) |
|  | Weeks 1-4 | 3.91 (3.76-4.07) | 1.52 (1.44-1.61) | 5.46 (4.86-6.13) |
|  | Weeks 5-28 | 1.57 (1.53-1.60) | 1.33 (1.27-1.39) | 2.54 (2.25-2.87) |
|  | Weeks 29-52 | 1.41 (1.37-1.44) | - | - |
|  | Weeks 53-102 | 1.64 (1.57-1.71) | - | - |
| Dyspepsia | Day 0 | 39.2 (36.1-42.6) | 8.14 (6.75-9.83) | 40.8 (31.3-53.1) |
|  | Weeks 1-4 | 1.92 (1.77-2.08) | 1.09 (0.97-1.22) | 3.01 (2.44-3.71) |
|  | Weeks 5-28 | 1.45 (1.40-1.51) | 1.23 (1.14-1.32) | 2.23 (1.86-2.68) |
|  | Weeks 29-52 | 1.43 (1.37-1.48) | - | - |
|  | Weeks 53-102 | 1.55 (1.44-1.67) | - | - |
| Peptic ulcer | Day 0 | 69.1 (57.7-82.8) | 16.2 (11.4-23.1) | 43.9 (20.2-95.4) |
|  | Weeks 1-4 | 4.87 (4.17-5.67) | 2.26 (1.83-2.80) | 3.80 (2.13-6.76) |
|  | Weeks 5-28 | 1.89 (1.72-2.08) | 1.27 (1.04-1.56) | 2.62 (1.57-4.38) |
|  | Weeks 29-52 | 1.42 (1.28-1.59) | - | - |
|  | Weeks 53-102 | 1.50 (1.24-1.81) | - | - |
| Non-alcoholic steatohepatitis | Day 0 | 191 (142-257) | 29.5 (16.2-53.7) | X |
|  | Weeks 1-4 | 6.18 (4.49-8.50) | 2.47 (1.60-3.82) | X |
|  | Weeks 5-28 | 2.25 (1.85-2.73) | 1.13 (0.71-1.79) | X |
|  | Weeks 29-52 | 1.40 (1.11-1.78) | - | - |
|  | Weeks 53-102 | 2.55 (1.85-3.52) | - | - |
| Gallstones | Day 0 | 116 (109-123) | 28.2 (25.4-31.3) | 59.6 (46.4-76.6) |
|  | Weeks 1-4 | 3.18 (2.95-3.42) | 1.49 (1.36-1.64) | 4.10 (3.34-5.05) |
|  | Weeks 5-28 | 1.68 (1.61-1.74) | 1.22 (1.13-1.32) | 2.65 (2.19-3.20) |
|  | Weeks 29-52 | 1.40 (1.34-1.47) | - | - |
|  | Weeks 53-102 | 1.66 (1.54-1.78) | - | - |
| Irritable bowel syndrome | Day 0 | 49.8 (45.9-54.1) | 13.2 (11.4-15.1) | 52.2 (41.9-65.1) |
|  | Weeks 1-4 | 2.48 (2.31-2.68) | 1.29 (1.17-1.42) | 4.76 (4.06-5.57) |
|  | Weeks 5-28 | 1.29 (1.24-1.34) | 1.25 (1.16-1.34) | 2.14 (1.ß2.55) |
|  | Weeks 29-52 | 1.28 (1.23-1.33) | - | - |
|  | Weeks 53-102 | 1.38 (1.29-1.49) | - | - |

#### Supplementary Table 4. Adjusted hazard ratios (95% CI) comparing the incidence of gastrointestinal disease events after COVID-19 with the incidence before or without COVID-19, in the pre-vaccination, vaccinated and unvaccinated cohorts, in hospitalised subgroup. Hazard ratios are maximally adjusted.

| Outcome | Time since COVID-19 | Pre-vaccination cohort | Vaccinated cohort | Unvaccinated cohort |
| --- | --- | --- | --- | --- |
| Upper gastrointestinal bleeding | Day 0 | 516 (475-560) | 391 (354-431) | 398 (302-524) |
|  | Weeks 1-4 | 20.1 (18.4-22.1) | 15.2 (13.4-17.3) | 19.7 (15.0-25.9) |
|  | Weeks 5-28 | 2.79 (2.52-3.09) | 3.78 (3.24-4.41) | 2.85 (1.80-4.53) |
|  | Weeks 29-52 | 1.78 (1.57-2.02) | - | - |
|  | Weeks 53-102 | 1.90 (1.63-2.22) | - | - |
| Lower gastrointestinal bleeding | Day 0 | 332 (302-366) | 160 (132-194) | 456 (383-543) |
|  | Weeks 1-4 | 13.7 (12.4-15.2) | 10.4 (8.80-12.2) | 11.4 (8.48-15.4) |
|  | Weeks 5-28 | 2.26 (2.04-2.51) | 3.40 (2.89-4.00) | 3.00 (2.23-4.04) |
|  | Weeks 29-52 | 1.51 (1.32-1.72) | - | - |
|  | Weeks 53-102 | 1.64 (1.39-1.93) | - | - |
| Gastro-oesophageal reflux | Day 0 | 373 (351-395) | 196 (176-218) | 396 (335-468) |
|  | Weeks 1-4 | 23.8 (22.6-25.1) | 15.2 (13.8-16.6) | 25.8 (22.2-29.9) |
|  | Weeks 5-28 | 2.10 (1.95-2.25) | 4.35 (3.88-4.87) | 2.89 (2.18-3.81) |
|  | Weeks 29-52 | 1.65 (1.53-1.78) | - | - |
|  | Weeks 53-102 | 1.65 (1.49-1.83) | - | - |
| Dyspepsia | Day 0 | 128 (108-152) | 73.0 (50.0-106) | 210 (158-280) |
|  | Weeks 1-4 | 7.03 (6.05-8.17) | 5.25 (3.90-7.07) | 9.83 (7.03-13.7) |
|  | Weeks 5-28 | 1.52 (1.33-1.73) | 1.92 (1.34-2.75) | 5.31 (4.21-6.69) |
|  | Weeks 29-52 | 1.56 (1.36-1.79) | - | - |
|  | Weeks 53-102 | 1.42 (1.16-1.73) | - | - |
| Gallstones | Day 0 | 261 (233-292) | 139 (115-169) | 176 (123-253) |
|  | Weeks 1-4 | 12.4 (11.1-13.9) | 9.53 (8.05-11.3) | 25.8 (21.5-31.1) |
|  | Weeks 5-28 | 2.36 (2.13-2.62) | 3.02 (2.53-3.61) | 2.91 (1.97-4.30) |
|  | Weeks 29-52 | 1.53 (1.34-1.74) | - | - |
|  | Weeks 53-102 | 1.83 (1.57-2.13) | - | - |
| Irritable bowel syndrome | Day 0 | 369 (327-417) | 232 (189-284) | 378 (290-493) |
|  | Weeks 1-4 | 23.0 (20.8-25.5) | 16.7 (13.8-20.3) | 27.3 (22.1-33.9) |
|  | Weeks 5-28 | 1.58 (1.36-1.84) | 3.52 (2.67-4.64) | 3.30 (2.23-4.88) |
|  | Weeks 29-52 | 1.42 (1.21-1.66) | - | - |
|  | Weeks 53-102 | 1.49 (1.21-1.84) | - | - |

X insufficient events

#### Supplementary Table 5. Adjusted hazard ratios (95% CI) comparing the incidence of gastrointestinal disease events after COVID-19 with the incidence before or without COVID-19, in the pre-vaccination, vaccinated and unvaccinated cohorts, in non-hospitalised subgroup. Hazard ratios are maximally adjusted.

| Outcome | Time since COVID-19 | Pre-vaccination cohort | Vaccinated cohort | Unvaccinated cohort |
| --- | --- | --- | --- | --- |
| Upper gastrointestinal bleeding | Day 0 | 131 (123-139) | 41.1 (37.1-45.6) | 61.7 (47.7-79.7) |
|  | Weeks 1-4 | 1.92 (1.74-2.11) | 1.20 (1.06-1.36) | 2.14 (1.61-2.85) |
|  | Weeks 5-28 | 1.46 (1.40-1.53) | 1.14 (1.04-1.25) | 1.15 (0.88-1.51) |
|  | Weeks 29-52 | 1.23 (1.18-1.29) | - | - |
|  | Weeks 53-102 | 1.34 (1.23-1.45) | - | - |
| Lower gastrointestinal bleeding | Day 0 | 65.7 (61.2-70.5) | 23.0 (20.5-25.8) | 31.2 (23.0-42.4) |
|  | Weeks 1-4 | 1.44 (1.32-1.59) | 1.02 (0.91-1.14) | 1.82 (1.39-2.38) |
|  | Weeks 5-28 | 1.39 (1.34-1.44) | 1.15 (1.07-1.24) | 1.28 (1.02-1.60) |
|  | Weeks 29-52 | 1.22 (1.17-1.27) | - | - |
|  | Weeks 53-102 | 1.30 (1.21-1.40) | - | - |
| Gastro-oesophageal reflux | Day 0 | 37.4 (35.2-39.9) | 11.8 (10.8-13.0) | 16.1 (12.3-21.2) |
|  | Weeks 1-4 | 1.46 (1.37-1.55) | 0.91 (0.84-0.97) | 1.40 (1.15-1.70) |
|  | Weeks 5-28 | 1.28 (1.25-1.32) | 1.14 (1.09-1.20) | 1.48 (1.29-1.70) |
|  | Weeks 29-52 | 1.17 (1.13-1.20) | - | - |
|  | Weeks 53-102 | 1.28 (1.22-1.34) | - | - |
| Dyspepsia | Day 0 | 20.3 (18.0-22.9) | 5.37 (4.23-6.81) | 20.2 (14.4-28.4) |
|  | Weeks 1-4 | 1.25 (1.14-1.38) | 0.92 (0.81-1.04) | 1.52 (1.17-1.98) |
|  | Weeks 5-28 | 1.23 (1.19-1.28) | 1.10 (1.02-1.20) | 1.40 (1.15-1.72) |
|  | Weeks 29-52 | 1.20 (1.15-1.25) | - | - |
|  | Weeks 53-102 | 1.22 (1.13-1.32) | - | - |
| Gallstones | Day 0 | 78.0 (72.4-84.1) | 20.5 (18.1-23.1) | 27.2 (19.5-37.9) |
|  | Weeks 1-4 | 1.67 (1.51-1.85) | 0.97 (0.86-1.09) | 1.48 (1.09-2.02) |
|  | Weeks 5-28 | 1.36 (1.30-1.42) | 1.06 (0.98-1.14) | 1.69 (1.37-2.09) |
|  | Weeks 29-52 | 1.17 (1.12-1.23) | - | - |
|  | Weeks 53-102 | 1.25 (1.16-1.36) | - | - |
| Irritable bowel syndrome | Day 0 | 22.3 (19.7-25.1) | 7.47 (6.24-8.94) | 10.8 (7.13-16.2) |
|  | Weeks 1-4 | 1.08 (0.97-1.21) | 0.78 (0.69-0.87) | 1.45 (1.14-1.85) |
|  | Weeks 5-28 | 1.12 (1.07-1.16) | 1.05 (0.98-1.13) | 1.18 (0.97-1.43) |
|  | Weeks 29-52 | 1.10 (1.06-1.15) | - | - |
|  | Weeks 53-102 | 1.11 (1.03-1.20) | - | - |

X insufficient events

#### Supplementary Table 6. Adjusted hazard ratios (95% CI) comparing the incidence of gastrointestinal disease events after COVID-19 with the incidence before or without COVID-19, in the pre-vaccination, vaccinated and unvaccinated cohorts, in 18–39 age group. Hazard ratios are maximally adjusted.

| Outcome | Time since COVID-19 | Pre-vaccination cohort | Vaccinated cohort | Unvaccinated cohort |
| --- | --- | --- | --- | --- |
| Upper gastrointestinal bleeding | Day 0 | 20.3 (15.3-26.9) | 7.61 (4.50-12.9) | 37.9 (24.9-57.9) |
|  | Weeks 1-4 | 1.69 (1.40-2.04) | 1.26 (0.95-1.67) | 2.81 (2.01-3.92) |
|  | Weeks 5-28 | 1.25 (1.15-1.36) | 1.19 (0.97-1.47) | 1.51 (1.07-2.15) |
|  | Weeks 29-52 | 1.19 (1.08-1.30) | - | - |
|  | Weeks 53-102 | 1.35 (1.14-1.61) | - | - |
| Lower gastrointestinal bleeding | Day 0 | 8.91 (6.56-12.1) | 2.78 (1.45-5.36) | 11.4 (6.05-21.6) |
|  | Weeks 1-4 | 1.16 (0.99-1.37) | 0.88 (0.68-1.14) | 1.88 (1.34-2.65) |
|  | Weeks 5-28 | 1.27 (1.19-1.35) | 1.30 (1.12-1.51) | 1.49 (1.10-2.00) |
|  | Weeks 29-52 | 1.21 (1.13-1.30) | - | - |
|  | Weeks 53-102 | 1.40 (1.22-1.60) | - | - |
| Gastro-oesophageal reflux | Day 0 | 1.43 (1.28-1.60) | 0.85 (0.71-1.02) | 2.23 (1.81-2.76) |
|  | Weeks 1-4 | 1.19 (1.13-1.25) | 1.07 (0.95-1.20) | 1.64 (1.36-1.98) |
|  | Weeks 5-28 | 1.22 (1.16-1.28) | - | - |
|  | Weeks 29-52 | 1.38 (1.25-1.52) | - | - |
|  | Weeks 53-102 | 1.43 (1.28-1.60) | 0.85 (0.71-1.02) | 2.23 (1.81-2.76) |
| Dyspepsia | Day 0 | 12.6 (9.90-16.1) | 3.05 (1.64-5.67) | 20.9 (14.0-31.4) |
|  | Weeks 1-4 | 1.36 (1.18-1.58) | 0.87 (0.67-1.12) | 1.58 (1.15-2.17) |
|  | Weeks 5-28 | 1.20 (1.13-1.28) | 1.18 (1.01-1.38) | 1.51 (1.17-1.95) |
|  | Weeks 29-52 | 1.22 (1.14-1.30) | - | - |
|  | Weeks 53-102 | 1.24 (1.09-1.42) | - | - |
| Gallstones | Day 0 | 14.3 (10.4-19.6) | 3.51 (1.89-6.53) | 8.94 (4.43-18.0) |
|  | Weeks 1-4 | 1.33 (1.08-1.63) | 0.70 (0.52-0.95) | 1.33 (0.89-1.99) |
|  | Weeks 5-28 | 1.13 (1.04-1.23) | 0.96 (0.79-1.16) | 1.49 (1.11-2.02) |
|  | Weeks 29-52 | 1.12 (1.03-1.22) | - | - |
|  | Weeks 53-102 | 1.27 (1.08-1.51) | - | - |
| Irritable bowel syndrome | Day 0 | 10.7 (8.39-13.6) | 2.50 (1.48-4.23) | 10.2 (6.20-16.7) |
|  | Weeks 1-4 | 1.16 (1.01-1.34) | 0.89 (0.73-1.08) | 1.96 (1.54-2.50) |
|  | Weeks 5-28 | 1.10 (1.04-1.17) | 1.13 (1.00-1.28) | 1.11 (0.86-1.43) |
|  | Weeks 29-52 | 1.16 (1.09-1.23) | - | - |
|  | Weeks 53-102 | 1.20 (1.07-1.35) | - | - |

X insufficient events

#### Supplementary Table 7. Adjusted hazard ratios (95% CI) comparing the incidence of gastrointestinal disease events after COVID-19 with the incidence before or without COVID-19, in the pre-vaccination, vaccinated and unvaccinated cohorts, in 40–59 age group. Hazard ratios are maximally adjusted.

| Outcome | Time since COVID-19 | Pre-vaccination cohort | Vaccinated cohort | Unvaccinated cohort |
| --- | --- | --- | --- | --- |
| Upper gastrointestinal bleeding | Day 0 | 20.3 (15.3-26.9) | 7.61 (4.50-12.9) | 37.9 (24.9-57.9) |
|  | Weeks 1-4 | 1.69 (1.40-2.04) | 1.26 (0.95-1.67) | 2.81 (2.01-3.92) |
|  | Weeks 5-28 | 1.25 (1.15-1.36) | 1.19 (0.97-1.47) | 1.51 (1.07-2.15) |
|  | Weeks 29-52 | 1.19 (1.08-1.30) | - | - |
|  | Weeks 53-102 | 1.35 (1.14-1.61) | - | - |
| Lower gastrointestinal bleeding | Day 0 | 8.91 (6.56-12.1) | 2.78 (1.45-5.36) | 11.4 (6.05-21.6) |
|  | Weeks 1-4 | 1.16 (0.99-1.37) | 0.88 (0.68-1.14) | 1.88 (1.34-2.65) |
|  | Weeks 5-28 | 1.27 (1.19-1.35) | 1.30 (1.12-1.51) | 1.49 (1.10-2.00) |
|  | Weeks 29-52 | 1.21 (1.13-1.30) | - | - |
|  | Weeks 53-102 | 1.40 (1.22-1.60) | - | - |
| Gastro-oesophageal reflux | Day 0 | 1.43 (1.28-1.60) | 0.85 (0.71-1.02) | 2.23 (1.81-2.76) |
|  | Weeks 1-4 | 1.19 (1.13-1.25) | 1.07 (0.95-1.20) | 1.64 (1.36-1.98) |
|  | Weeks 5-28 | 1.22 (1.16-1.28) | - | - |
|  | Weeks 29-52 | 1.38 (1.25-1.52) | - | - |
|  | Weeks 53-102 | 1.43 (1.28-1.60) | 0.85 (0.71-1.02) | 2.23 (1.81-2.76) |
| Dyspepsia | Day 0 | 12.6 (9.90-16.1) | 3.05 (1.64-5.67) | 20.9 (14.0-31.4) |
|  | Weeks 1-4 | 1.36 (1.18-1.58) | 0.87 (0.67-1.12) | 1.58 (1.15-2.17) |
|  | Weeks 5-28 | 1.20 (1.13-1.28) | 1.18 (1.01-1.38) | 1.51 (1.17-1.95) |
|  | Weeks 29-52 | 1.22 (1.14-1.30) | - | - |
|  | Weeks 53-102 | 1.24 (1.09-1.42) | - | - |
| Gallstones | Day 0 | 14.3 (10.4-19.6) | 3.51 (1.89-6.53) | 8.94 (4.43-18.0) |
|  | Weeks 1-4 | 1.33 (1.08-1.63) | 0.70 (0.52-0.95) | 1.33 (0.89-1.99) |
|  | Weeks 5-28 | 1.13 (1.04-1.23) | 0.96 (0.79-1.16) | 1.49 (1.11-2.02) |
|  | Weeks 29-52 | 1.12 (1.03-1.22) | - | - |
|  | Weeks 53-102 | 1.27 (1.08-1.51) | - | - |
| Irritable bowel syndrome | Day 0 | 10.7 (8.39-13.6) | 2.50 (1.48-4.23) | 10.2 (6.20-16.7) |
|  | Weeks 1-4 | 1.16 (1.01-1.34) | 0.89 (0.73-1.08) | 1.96 (1.54-2.50) |
|  | Weeks 5-28 | 1.10 (1.04-1.17) | 1.13 (1.00-1.28) | 1.11 (0.86-1.43) |
|  | Weeks 29-52 | 1.16 (1.09-1.23) | - | - |
|  | Weeks 53-102 | 1.20 (1.07-1.35) | - | - |

#### X insufficient events

#### Supplementary Table 8. Adjusted hazard ratios (95% CI) comparing the incidence of gastrointestinal disease events after COVID-19 with the incidence before or without COVID-19, in the pre-vaccination, vaccinated and unvaccinated cohorts, in 60–79 age group. Hazard ratios are maximally adjusted.

| Outcome | Time since COVID-19 | Pre-vaccination cohort | Vaccinated cohort | Unvaccinated cohort |
| --- | --- | --- | --- | --- |
| Upper gastrointestinal bleeding | Day 0 | 322 (299-347) | 70.2 (60.9-81.0) | 382 (278-524) |
|  | Weeks 1-4 | 13.7 (12.9-14.5) | 2.23 (1.90-2.62) | 8.78 (5.71-13.5) |
|  | Weeks 5-28 | 2.40 (2.25-2.56) | 1.19 (1.02-1.39) | 1.17 (0.55-2.48) |
|  | Weeks 29-52 | 1.41 (1.29-1.53) | - | - |
|  | Weeks 53-102 | 2.00 (1.81-2.22) | - | - |
| Lower gastrointestinal bleeding | Day 0 | 192 (176-210) | 48.5 (41.6-56.4) | 260 (181-373) |
|  | Weeks 1-4 | 4.75 (4.24-5.31) | 1.92 (1.63-2.25) | 13.0 (9.86-17.1) |
|  | Weeks 5-28 | 1.70 (1.58-1.83) | 1.14 (0.98-1.32) | 1.39 (0.73-2.64) |
|  | Weeks 29-52 | 1.26 (1.16-1.38) | - | - |
|  | Weeks 53-102 | 1.54 (1.35-1.75) | - | - |
| Gastro-oesophageal reflux | Day 0 | 131 (123-140) | 25.7 (22.9-28.9) | 243 (194-305) |
|  | Weeks 1-4 | 5.68 (5.33-6.05) | 1.83 (1.67-2.00) | 13.3 (10.8-16.5) |
|  | Weeks 5-28 | 1.52 (1.45-1.59) | 1.28 (1.19-1.38) | 2.65 (2.02-3.48) |
|  | Weeks 29-52 | 1.20 (1.15-1.27) | - | - |
|  | Weeks 53-102 | 1.37 (1.27-1.48) | - | - |
| Dyspepsia | Day 0 | 51.4 (43.4-60.7) | 12.6 (9.32-17.1) | X |
|  | Weeks 1-4 | 2.35 (2.01-2.76) | 1.20 (0.98-1.49) | X |
|  | Weeks 5-28 | 1.30 (1.20-1.42) | 1.13 (0.97-1.32) | X |
|  | Weeks 29-52 | 1.21 (1.10-1.32) | - | - |
|  | Weeks 53-102 | 1.26 (1.08-1.46) | - | - |
| Gallstones | Day 0 | 182 (166-200) | 40.7 (34.7-47.7) | 285 (209-388) |
|  | Weeks 1-4 | 4.29 (3.79-4.85) | 1.91 (1.65-2.23) | 13.2 (9.93-17.7) |
|  | Weeks 5-28 | 1.73 (1.61-1.87) | 1.16 (1.01-1.33) | 3.08 (2.01-4.70) |
|  | Weeks 29-52 | 1.31 (1.21-1.42) | - | - |
|  | Weeks 53-102 | 1.35 (1.18-1.54) | - | - |
| Irritable bowel syndrome | Day 0 | 122 (106-140) | 28.4 (22.8-35.3) | 342 (248-472) |
|  | Weeks 1-4 | 5.13 (4.47-5.89) | 1.57 (1.30-1.90) | 15.1 (10.5-21.9) |
|  | Weeks 5-28 | 1.24 (1.12-1.38) | 1.12 (0.96-1.31) | 7.68 (5.65-10.5) |
|  | Weeks 29-52 | 1.13 (1.02-1.25) | - | - |
|  | Weeks 53-102 | 1.00 (0.84-1.19) | - | - |

#### X insufficient events

#### Supplementary Table 9. Adjusted hazard ratios (95% CI) comparing the incidence of gastrointestinal disease events after COVID-19 with the incidence before or without COVID-19, in the pre-vaccination, vaccinated and unvaccinated cohorts, in 80–110 age group. Hazard ratios are maximally adjusted.

| Outcome | Time since COVID-19 | Pre-vaccination cohort | Vaccinated cohort | Unvaccinated cohort |
| --- | --- | --- | --- | --- |
| Upper gastrointestinal bleeding | Day 0 | 398 (366-432) | 213 (187-242) | - |
|  | Weeks 1-4 | 4.86 (4.18-5.65) | 4.39 (3.64-5.30) | - |
|  | Weeks 5-28 | 2.02 (1.83-2.24) | 1.37 (1.08-1.73) | - |
|  | Weeks 29-52 | 1.41 (1.24-1.59) | - | - |
|  | Weeks 53-102 | 1.31 (1.12-1.52) | - | - |
| Lower gastrointestinal bleeding | Day 0 | 316 (287-348) | 196 (170-226) | - |
|  | Weeks 1-4 | 4.82 (4.11-5.66) | 3.71 (3.00-4.59) | - |
|  | Weeks 5-28 | 1.85 (1.65-2.07) | 1.21 (0.93-1.57) | - |
|  | Weeks 29-52 | 1.31 (1.14-1.50) | - | - |
|  | Weeks 53-102 | 1.23 (1.04-1.46) | - | - |
| Gastro-oesophageal reflux | Day 0 | 399 (376-424) | 123 (109-138) | - |
|  | Weeks 1-4 | 12.2 (11.5-13.0) | 3.72 (3.23-4.29) | - |
|  | Weeks 5-28 | 1.94 (1.79-2.10) | 1.55 (1.33-1.80) | - |
|  | Weeks 29-52 | 1.47 (1.34-1.62) | - | - |
|  | Weeks 53-102 | 1.28 (1.14-1.44) | - | - |
| Dyspepsia | Day 0 | 98.1 (77.5-124) | 65.3 (46.6-91.5) | - |
|  | Weeks 1-4 | 2.39 (1.73-3.29) | 2.84 (2.01-4.03) | - |
|  | Weeks 5-28 | 1.45 (1.21-1.74) | 1.65 (1.19-2.29) | - |
|  | Weeks 29-52 | 1.16 (0.93-1.44) | - | - |
|  | Weeks 53-102 | 1.14 (0.87-1.48) | - | - |
| Gallstones | Day 0 | 252 (226-281) | 135 (115-159) | - |
|  | Weeks 1-4 | 4.29 (3.62-5.09) | 2.68 (2.11-3.40) | - |
|  | Weeks 5-28 | 1.81 (1.62-2.03) | 1.57 (1.26-1.97) | - |
|  | Weeks 29-52 | 1.16 (1.00-1.34) | - | - |
|  | Weeks 53-102 | 1.20 (1.02-1.42) | - | - |
| Irritable bowel syndrome | Day 0 | 284 (235-342) | 109 (82.6-145) | - |
|  | Weeks 1-4 | 6.22 (4.80-8.06) | 3.73 (2.72-5.10) | - |
|  | Weeks 5-28 | 1.56 (1.27-1.91) | 1.45 (1.02-2.06) | - |
|  | Weeks 29-52 | 1.31 (1.06-1.62) | - | - |
|  | Weeks 53-102 | 1.38 (1.08-1.75) | - | - |

X insufficient events

#### Supplementary Table 10 Adjusted hazard ratios (95% CI) comparing the incidence of gastrointestinal disease events after COVID-19 with the incidence before or without COVID-19, in the pre-vaccination, vaccinated and unvaccinated cohorts, in White ethnicity. Hazard ratios are maximally adjusted.

| Subgroup and outcome | Time since COVID-19 | Pre-vaccination cohort | Vaccinated cohort | Unvaccinated cohort |
| --- | --- | --- | --- | --- |
| Upper gastrointestinal bleeding | Day 0 | 187 (177-197) | 50.9 (46.3-55.9) | 94.9 (76.9-117) |
|  | Weeks 1-4 | 3.53 (3.28-3.80) | 1.83 (1.65-2.02) | 3.46 (2.75-4.35) |
|  | Weeks 5-28 | 1.57 (1.50-1.64) | 1.18 (1.08-1.29) | 1.26 (0.97-1.65) |
|  | Weeks 29-52 | 1.28 (1.22-1.34) | - | - |
|  | Weeks 53-102 | 1.42 (1.31-1.53) | - | - |
| Lower gastrointestinal bleeding | Day 0 | 96.3 (90.6-102) | 29.2 (26.3-32.4) | 52.9 (41.6-67.4) |
|  | Weeks 1-4 | 2.45 (2.27-2.64) | 1.42 (1.29-1.57) | 2.45 (1.93-3.10) |
|  | Weeks 5-28 | 1.43 (1.38-1.49) | 1.18 (1.09-1.27) | 1.17 (0.92-1.49) |
|  | Weeks 29-52 | 1.25 (1.20-1.31) | - | - |
|  | Weeks 53-102 | 1.31 (1.22-1.42) | - | - |
|  | Day 0 | 75.0 (71.5-78.6) | 18.4 (17.0-20.0) | 47.5 (40.0-56.3) |
| Gastro-oesophageal reflux | Weeks 1-4 | 3.26 (3.12-3.41) | 1.43 (1.35-1.52) | 3.55 (3.10-4.06) |
|  | Weeks 5-28 | 1.35 (1.32-1.39) | 1.19 (1.14-1.25) | 1.59 (1.38-1.84) |
|  | Weeks 29-52 | 1.18 (1.15-1.22) | - | - |
|  | Weeks 53-102 | 1.34 (1.28-1.41) | - | - |
| Dyspepsia | Day 0 | 30.0 (26.9-33.5) | 8.42 (6.96-10.2) | 21.3 (14.9-30.4) |
|  | Weeks 1-4 | 1.50 (1.36-1.65) | 1.05 (0.93-1.18) | 1.95 (1.52-2.51) |
|  | Weeks 5-28 | 1.24 (1.19-1.30) | 1.13 (1.04-1.23) | 1.41 (1.13-1.75) |
|  | Weeks 29-52 | 1.20 (1.15-1.26) | - | - |
|  | Weeks 53-102 | 1.23 (1.13-1.33) | - | - |
| Gallstones | Day 0 | 104 (97.7-111) | 26.6 (23.9-29.7) | 37.2 (27.9-49.7) |
|  | Weeks 1-4 | 2.73 (2.52-2.96) | 1.39 (1.26-1.54) | 2.77 (2.20-3.50) |
|  | Weeks 5-28 | 1.49 (1.42-1.55) | 1.10 (1.01-1.19) | 1.73 (1.40-2.14) |
|  | Weeks 29-52 | 1.20 (1.15-1.26) | - | - |
|  | Weeks 53-102 | 1.34 (1.24-1.45) | - | - |
| Irritable bowel syndrome | Day 0 | 44.3 (40.5-48.5) | 13.8 (12.1-15.7) | 32.6 (25.6-41.5) |
|  | Weeks 1-4 | 2.18 (2.01-2.36) | 1.16 (1.05-1.28) | 2.92 (2.45-3.48) |
|  | Weeks 5-28 | 1.15 (1.10-1.20) | 1.09 (1.01-1.17) | 1.35 (1.12-1.63) |
|  | Weeks 29-52 | 1.11 (1.06-1.16) | - | - |
|  | Weeks 53-102 | 1.14 (1.05-1.23) | - | - |

#### X insufficient events

#### Supplementary Table 11: Adjusted hazard ratios (95% CI) comparing the incidence of gastrointestinal disease events after COVID-19 with the incidence before or without COVID-19, in the pre-vaccination, vaccinated and unvaccinated cohorts, in South Asian ethnicity. Hazard ratios are maximally adjusted.

| Subgroup and outcome | Time since COVID-19 | Pre-vaccination cohort | Vaccinated cohort | Unvaccinated cohort |
| --- | --- | --- | --- | --- |
| Upper gastrointestinal bleeding | Day 0 | 120 (96.3-149) | 72.7 (49.0-108) | X |
|  | Weeks 1-4 | 5.87 (4.83-7.14) | 2.01 (1.24-3.25) | X |
|  | Weeks 5-28 | 1.43 (1.23-1.67) | 1.36 (0.91-2.02) | X |
|  | Weeks 29-52 | 1.32 (1.13-1.54) | - | - |
|  | Weeks 53-102 | 1.36 (1.05-1.76) | - | - |
| Lower gastrointestinal bleeding | Day 0 | 57.4 (45.2-72.9) | 37.1 (24.2-57.0) | X |
|  | Weeks 1-4 | 2.83 (2.29-3.51) | 1.02 (0.59-1.77) | X |
|  | Weeks 5-28 | 1.45 (1.29-1.63) | 1.16 (0.81-1.65) | X |
|  | Weeks 29-52 | 1.21 (1.06-1.38) | - | - |
|  | Weeks 53-102 | 1.53 (1.25-1.88) | - | - |
|  | Day 0 | 54.1 (47.0-62.3) | 20.7 (15.3-28.2) | 127 (90.4-177) |
| Gastro-oesophageal reflux | Weeks 1-4 | 3.81 (3.42-4.23) | 1.33 (1.04-1.72) | 6.38 (4.64-8.77) |
|  | Weeks 5-28 | 1.26 (1.18-1.36) | 1.21 (1.01-1.45) | 2.18 (1.53-3.10) |
|  | Weeks 29-52 | 1.22 (1.14-1.31) | - | - |
|  | Weeks 53-102 | 1.25 (1.11-1.41) | - | - |
| Dyspepsia | Day 0 | 21.4 (16.0-28.5) | 7.05 (3.17-15.7) | X |
|  | Weeks 1-4 | 2.12 (1.77-2.54) | 0.96 (0.61-1.51) | X |
|  | Weeks 5-28 | 1.31 (1.19-1.43) | 1.00 (0.74-1.35) | X |
|  | Weeks 29-52 | 1.25 (1.13-1.38) | - | - |
|  | Weeks 53-102 | 1.31 (1.10-1.56) | - | - |
| Gallstones | Day 0 | 56.3 (42.9-73.9) | 16.8 (8.69-32.3) | X |
|  | Weeks 1-4 | 2.59 (2.02-3.33) | 1.53 (0.97-2.40) | X |
|  | Weeks 5-28 | 1.27 (1.10-1.45) | 0.91 (0.61-1.35) | X |
|  | Weeks 29-52 | 1.17 (1.01-1.35) | - | - |
|  | Weeks 53-102 | 1.36 (1.08-1.70) | - | - |
| Irritable bowel syndrome | Day 0 | 30.1 (20.9-43.3) | X | X |
|  | Weeks 1-4 | 2.32 (1.79-3.01) | X | X |
|  | Weeks 5-28 | 1.08 (0.93-1.25) | X | X |
|  | Weeks 29-52 | 1.13 (0.98-1.31) | - | - |
|  | Weeks 53-102 | 1.10 (0.85-1.42) | - | - |

X insufficient events

#### Supplementary Table 12: Adjusted hazard ratios (95% CI) comparing the incidence of gastrointestinal disease events after COVID-19 with the incidence before or without COVID-19, in the pre-vaccination, vaccinated and unvaccinated cohorts, in Female sex. Hazard ratios are maximally adjusted.

| Subgroup and outcome | Time since COVID-19 | Pre-vaccination cohort | Vaccinated cohort | Unvaccinated cohort |
| --- | --- | --- | --- | --- |
| Upper gastrointestinal bleeding | Day 0 | 148 (137-160) | 44.0 (38.4-50.4) | 98.4 (75.3-129) |
|  | Weeks 1-4 | 2.81 (2.53-3.13) | 1.62 (1.40-1.87) | 4.27 (3.26-5.58) |
|  | Weeks 5-28 | 1.48 (1.39-1.56) | 1.23 (1.10-1.39) | 1.22 (0.86-1.72) |
|  | Weeks 29-52 | 1.24 (1.17-1.33) | - | - |
|  | Weeks 53-102 | 1.36 (1.23-1.50) | - | - |
| Lower gastrointestinal bleeding | Day 0 | 76.1 (69.7-83.0) | 23.5 (20.2-27.4) | 50.4 (36.9-68.8) |
|  | Weeks 1-4 | 1.89 (1.70-2.11) | 1.20 (1.04-1.38) | 2.62 (1.97-3.50) |
|  | Weeks 5-28 | 1.35 (1.28-1.42) | 1.22 (1.11-1.35) | 1.18 (0.88-1.60) |
|  | Weeks 29-52 | 1.20 (1.13-1.27) | - | - |
|  | Weeks 53-102 | 1.25 (1.14-1.38) | - | - |
| Gastro-oesophageal reflux | Day 0 | 58.5 (54.9-62.2) | 18.7 (17.1-20.5) | 46.8 (38.5-56.8) |
|  | Weeks 1-4 | 2.90 (2.75-3.07) | 1.23 (1.13-1.33) | 3.12 (2.65-3.67) |
|  | Weeks 5-28 | 1.29 (1.25-1.33) | 1.16 (1.10-1.23) | 1.51 (1.28-1.77) |
|  | Weeks 29-52 | 1.17 (1.14-1.21) | - | - |
|  | Weeks 53-102 | 1.29 (1.22-1.36) | - | - |
| Dyspepsia | Day 0 | 24.6 (21.5-28.0) | 7.51 (5.92-9.52) | 30.7 (22.4-42.2) |
|  | Weeks 1-4 | 1.60 (1.45-1.78) | 0.94 (0.81-1.08) | 2.03 (1.56-2.64) |
|  | Weeks 5-28 | 1.24 (1.18-1.29) | 1.11 (1.01-1.23) | 1.58 (1.27-1.97) |
|  | Weeks 29-52 | 1.20 (1.14-1.26) | - | - |
|  | Weeks 53-102 | 1.23 (1.13-1.35) | - | - |
| Gallstones | Day 0 | 75.1 (68.9-81.8) | 20.2 (17.5-23.4) | 35.1 (26.0-47.4) |
|  | Weeks 1-4 | 2.19 (1.98-2.42) | 1.20 (1.06-1.36) | 2.06 (1.57-2.69) |
|  | Weeks 5-28 | 1.35 (1.29-1.42) | 1.07 (0.97-1.17) | 1.69 (1.36-2.10) |
|  | Weeks 29-52 | 1.17 (1.11-1.24) | - | - |
|  | Weeks 53-102 | 1.31 (1.21-1.43) | - | - |
| Irritable bowel syndrome | Day 0 | 38.3 (34.5-42.6) | 11.8 (10.0-13.8) | 32.2 (24.8-41.7) |
|  | Weeks 1-4 | 2.04 (1.86-2.23) | 1.10 (0.98-1.23) | 3.12 (2.61-3.74) |
|  | Weeks 5-28 | 1.11 (1.06-1.16) | 1.12 (1.03-1.21) | 1.38 (1.14-1.68) |
|  | Weeks 29-52 | 1.10 (1.05-1.15) | - | - |
|  | Weeks 53-102 | 1.12 (1.03-1.22) | - | - |

X insufficient events

#### Supplementary Table 13: Adjusted hazard ratios (95% CI) comparing the incidence of gastrointestinal disease events after COVID-19 with the incidence before or without COVID-19, in the pre-vaccination, vaccinated and unvaccinated cohorts, in Male sex. Hazard ratios are maximally adjusted.

| Subgroup and outcome | Time since COVID-19 | Pre-vaccination cohort | Vaccinated cohort | Unvaccinated cohort |
| --- | --- | --- | --- | --- |
| Upper gastrointestinal bleeding | Day 0 | 216 (202-231) | 64.4 (57.5-72.1) | 114 (87.3-148) |
|  | Weeks 1-4 | 5.04 (4.63-5.49) | 2.10 (1.83-2.39) | 4.22 (3.17-5.61) |
|  | Weeks 5-28 | 1.72 (1.62-1.82) | 1.17 (1.03-1.32) | 1.60 (1.16-2.21) |
|  | Weeks 29-52 | 1.34 (1.26-1.43) | - | - |
|  | Weeks 53-102 | 1.53 (1.38-1.70) | - | - |
| Lower gastrointestinal bleeding | Day 0 | 110 (102-119) | 36.6 (32.1-41.7) | 64.2 (47.0-87.6) |
|  | Weeks 1-4 | 3.17 (2.90-3.47) | 1.66 (1.46-1.89) | 3.23 (2.41-4.34) |
|  | Weeks 5-28 | 1.58 (1.50-1.67) | 1.16 (1.04-1.29) | 1.61 (1.20-2.15) |
|  | Weeks 29-52 | 1.30 (1.22-1.37) | - | - |
|  | Weeks 53-102 | 1.46 (1.32-1.61) | - | - |
| Gastro-oesophageal reflux | Day 0 | 88.9 (83.6-94.5) | 22.5 (20.2-25.0) | 71.8 (58.4-88.3) |
|  | Weeks 1-4 | 4.05 (3.83-4.29) | 1.71 (1.58-1.85) | 5.06 (4.27-6.00) |
|  | Weeks 5-28 | 1.42 (1.37-1.48) | 1.26 (1.18-1.35) | 1.94 (1.60-2.34) |
|  | Weeks 29-52 | 1.23 (1.18-1.28) | - | - |
|  | Weeks 53-102 | 1.36 (1.27-1.46) | - | - |
| Dyspepsia | Day 0 | 35.8 (30.7-41.6) | 8.22 (6.02-11.2) | 32.3 (20.2-51.6) |
|  | Weeks 1-4 | 1.73 (1.51-1.98) | 1.21 (1.01-1.45) | 2.75 (1.94-3.90) |
|  | Weeks 5-28 | 1.28 (1.20-1.36) | 1.14 (1.00-1.30) | 1.53 (1.09-2.14) |
|  | Weeks 29-52 | 1.26 (1.17-1.34) | - | - |
|  | Weeks 53-102 | 1.27 (1.12-1.44) | - | - |
| Gallstones | Day 0 | 149 (136-164) | 39.8 (34.1-46.5) | 80.0 (50.4-127) |
|  | Weeks 1-4 | 3.97 (3.54-4.44) | 1.75 (1.50-2.04) | 7.00 (4.93-9.93) |
|  | Weeks 5-28 | 1.64 (1.53-1.76) | 1.14 (0.99-1.30) | 1.84 (1.17-2.90) |
|  | Weeks 29-52 | 1.26 (1.17-1.37) | - | - |
|  | Weeks 53-102 | 1.40 (1.23-1.58) | - | - |
| Irritable bowel syndrome | Day 0 | 53.7 (46.0-62.6) | 16.2 (12.7-20.8) | 54.9 (36.4-83.0) |
|  | Weeks 1-4 | 2.75 (2.41-3.15) | 1.48 (1.24-1.77) | 3.98 (2.87-5.51) |
|  | Weeks 5-28 | 1.24 (1.15-1.34) | 1.05 (0.90-1.22) | 1.24 (0.82-1.88) |
|  | Weeks 29-52 | 1.15 (1.06-1.25) | - | - |
|  | Weeks 53-102 | 1.13 (0.97-1.31) | - | - |

X insufficient events

#### Supplementary Table 14: Adjusted hazard ratios (95% CI) comparing the incidence of gastrointestinal disease events after COVID-19 with the incidence before or without COVID-19, in the pre-vaccination, vaccinated and unvaccinated cohorts, in individuals with prior history of gastrointestinal diseases. Hazard ratios are maximally adjusted.

| Outcome | Time since COVID-19 | Pre-vaccination cohort | Vaccinated cohort | Unvaccinated cohort |
| --- | --- | --- | --- | --- |
| Upper gastrointestinal bleeding | Day 0 | 160 (151-170) | 47.4 (42.6-52.7) | 84.1 (66.8-106) |
|  | Weeks 1-4 | 3.36 (3.09-3.64) | 1.84 (1.64-2.05) | 3.00 (2.33-3.88) |
|  | Weeks 5-28 | 1.53 (1.46-1.61) | 1.17 (1.06-1.29) | 1.25 (0.95-1.64) |
|  | Weeks 29-52 | 1.25 (1.18-1.32) | - | - |
|  | Weeks 53-102 | 1.47 (1.35-1.60) | - | - |
| Lower gastrointestinal bleeding | Day 0 | 83.7 (78.0-89.8) | 27.6 (24.5-31.1) | 44.3 (33.7-58.2) |
|  | Weeks 1-4 | 2.31 (2.13-2.51) | 1.42 (1.27-1.58) | 2.20 (1.69-2.85) |
|  | Weeks 5-28 | 1.41 (1.35-1.47) | 1.14 (1.05-1.24) | 1.22 (0.95-1.56) |
|  | Weeks 29-52 | 1.19 (1.14-1.25) | - | - |
|  | Weeks 53-102 | 1.34 (1.24-1.45) | - | - |
| Gastro-oesophageal reflux | Weeks 1-4 | 76.3 (72.8-79.9) | 19.8 (18.3-21.4) | 56.8 (48.8-66.2) |
|  | Weeks 5-28 | 3.49 (3.34-3.64) | 1.46 (1.37-1.55) | 3.78 (3.33-4.29) |
|  | Weeks 29-52 | 1.34 (1.30-1.37) | 1.20 (1.15-1.26) | 1.56 (1.36-1.79) |
|  | Weeks 53-102 | 1.19 (1.15-1.22) | - | - |
|  | Weeks 1-4 | 1.31 (1.25-1.37) | - | - |
| Dyspepsia | Day 0 | 26.7 (23.8-29.9) | 9.39 (7.80-11.3) | 26.0 (18.9-35.7) |
|  | Weeks 1-4 | 1.56 (1.42-1.72) | 1.02 (0.90-1.16) | 1.91 (1.49-2.46) |
|  | Weeks 5-28 | 1.22 (1.17-1.27) | 1.11 (1.02-1.21) | 1.44 (1.17-1.78) |
|  | Weeks 29-52 | 1.19 (1.14-1.25) | - | - |
|  | Weeks 53-102 | 1.25 (1.15-1.36) | - | - |
| Gallstones | Day 0 | 94.6 (87.9-102) | 24.4 (21.6-27.5) | 33.2 (24.6-44.9) |
|  | Weeks 1-4 | 2.75 (2.53-3.00) | 1.37 (1.23-1.52) | 2.35 (1.84-3.00) |
|  | Weeks 5-28 | 1.40 (1.34-1.47) | 1.08 (0.99-1.17) | 1.29 (1.02-1.62) |
|  | Weeks 29-52 | 1.19 (1.14-1.26) | - | - |
|  | Weeks 53-102 | 94.6 (87.9-102) | 24.4 (21.6-27.5) | 33.2 (24.6-44.9) |
| Irritable bowel syndrome | Day 0 | 46.1 (42.1-50.4) | 13.2 (11.5-15.2) | 36.5 (29.0-46.0) |
|  | Weeks 1-4 | 2.41 (2.23-2.60) | 1.21 (1.09-1.33) | 3.44 (2.92-4.05) |
|  | Weeks 5-28 | 1.11 (1.06-1.15) | 1.09 (1.01-1.17) | 1.36 (1.13-1.64) |
|  | Weeks 29-52 | 1.09 (1.04-1.14) | - | - |
|  | Weeks 53-102 | 1.13 (1.05-1.22) | - | - |

X insufficient events

#### Supplementary Table 15: Adjusted hazard ratios (95% CI) comparing the incidence of gastrointestinal disease events after COVID-19 with the incidence before or without COVID-19, in the pre-vaccination, vaccinated and unvaccinated cohorts, in individuals without prior history of gastrointestinal diseases. Hazard ratios are maximally adjusted.

| Subgroup and outcome | Time since COVID-19 | Pre-vaccination cohort | Vaccinated cohort | Unvaccinated cohort |
| --- | --- | --- | --- | --- |
| Upper gastrointestinal bleeding | Day 0 | 243 (222-265) | 72.0 (61.4-84.4) | 183 (133-252) |
|  | Weeks 1-4 | 5.36 (4.78-6.01) | 1.86 (1.51-2.28) | 8.98 (6.63-12.2) |
|  | Weeks 5-28 | 1.74 (1.61-1.88) | 1.30 (1.09-1.55) | 1.92 (1.22-3.00) |
|  | Weeks 29-52 | 1.40 (1.28-1.52) | - | - |
|  | Weeks 53-102 | 1.36 (1.16-1.58) | - | - |
| Lower gastrointestinal bleeding | Day 0 | 116 (105-129) | 34.6 (28.7-41.8) | 102 (71.0-147) |
|  | Weeks 1-4 | 3.00 (2.65-3.40) | 1.40 (1.15-1.70) | 5.53 (3.96-7.71) |
|  | Weeks 5-28 | 1.54 (1.44-1.65) | 1.35 (1.17-1.56) | 1.91 (1.29-2.85) |
|  | Weeks 29-52 | 1.36 (1.26-1.46) | - | - |
|  | Weeks 53-102 | 1.34 (1.17-1.54) | - | - |
| Gastro-oesophageal reflux | Weeks 1-4 | 47.4 (42.0-53.5) | 15.3 (12.7-18.4) | 49.9 (34.2-72.8) |
|  | Weeks 5-28 | 2.82 (2.56-3.12) | 1.24 (1.06-1.45) | 3.92 (2.93-5.26) |
|  | Weeks 29-52 | 1.38 (1.31-1.46) | 1.19 (1.07-1.34) | 2.17 (1.64-2.88) |
|  | Weeks 53-102 | 1.26 (1.19-1.33) | - | - |
|  | Weeks 1-4 | 1.42 (1.28-1.57) | - | - |
| Dyspepsia | Day 0 | 34.2 (28.2-41.5) | 3.83 (2.06-7.13) | 50.6 (31.2-82.1) |
|  | Weeks 1-4 | 1.92 (1.63-2.25) | 1.08 (0.84-1.39) | 3.65 (2.48-5.38) |
|  | Weeks 5-28 | 1.34 (1.24-1.45) | 1.18 (0.99-1.40) | 2.03 (1.39-2.98) |
|  | Weeks 29-52 | 1.31 (1.21-1.42) | - | - |
|  | Weeks 53-102 | 1.23 (1.05-1.44) | - | - |
| Gallstones | Day 0 | 117 (104-133) | 35.3 (28.4-43.9) | 88.1 (55.1-141) |
|  | Weeks 1-4 | 2.77 (2.37-3.23) | 1.45 (1.16-1.82) | 5.61 (3.76-8.36) |
|  | Weeks 5-28 | 1.55 (1.43-1.67) | 1.20 (1.00-1.42) | 3.97 (2.80-5.61) |
|  | Weeks 29-52 | 1.20 (1.10-1.31) | - | - |
|  | Weeks 53-102 | 1.24 (1.06-1.45) | - | - |
| Irritable bowel syndrome | Day 0 | 19.5 (14.0-27.2) | 3.54 (1.59-7.88) | X |
|  | Weeks 1-4 | 1.13 (0.86-1.48) | 0.88 (0.62-1.24) | X |
|  | Weeks 5-28 | 1.34 (1.22-1.48) | 1.09 (0.87-1.37) | X |
|  | Weeks 29-52 | 1.29 (1.17-1.42) | - | - |
|  | Weeks 53-102 | 1.08 (0.86-1.34) | - | - |

X insufficient events

#### Supplementary Table 16: Adjusted hazard ratios (95% CI) comparing the incidence of gastrointestinal disease events after COVID-19 with the incidence before or without COVID-19, in the pre-vaccination, vaccinated and unvaccinated cohorts, in individuals with prior history of gastrointestinal operations. Hazard ratios are maximally adjusted.

| Outcome | Time since COVID-19 | Pre-vaccination cohort | Vaccinated cohort | Unvaccinated cohort |
| --- | --- | --- | --- | --- |
| Upper gastrointestinal bleeding | Day 0 | 158 (144-174) | 42.1 (35.9-49.3) | 58.8 (38.8-89.0) |
|  | Weeks 1-4 | 3.10 (2.71-3.54) | 1.74 (1.49-2.05) | 2.54 (1.66-3.90) |
|  | Weeks 5-28 | 1.57 (1.45-1.69) | 1.11 (0.97-1.28) | 0.69 (0.40-1.21) |
|  | Weeks 29-52 | 1.32 (1.21-1.43) | - | - |
|  | Weeks 53-102 | 1.57 (1.38-1.78) | - | - |
| Lower gastrointestinal bleeding | Day 0 | 94.6 (84.8-106) | 26.4 (22.0-31.7) | 42.8 (27.5-66.6) |
|  | Weeks 1-4 | 2.34 (2.04-2.69) | 1.43 (1.21-1.68) | 1.98 (1.28-3.06) |
|  | Weeks 5-28 | 1.43 (1.33-1.53) | 1.10 (0.96-1.25) | 0.87 (0.55-1.37) |
|  | Weeks 29-52 | 1.20 (1.11-1.30) | - | - |
|  | Weeks 53-102 | 1.31 (1.15-1.49) | - | - |
| Gastro-oesophageal reflux | Weeks 1-4 | 83.4 (77.3-90.1) | 19.9 (17.7-22.4) | 53.5 (41.5-69.0) |
|  | Weeks 5-28 | 3.71 (3.45-3.99) | 1.48 (1.35-1.62) | 3.59 (2.90-4.43) |
|  | Weeks 29-52 | 1.31 (1.25-1.37) | 1.14 (1.06-1.23) | 1.64 (1.32-2.03) |
|  | Weeks 53-102 | 1.15 (1.10-1.21) | - | - |
|  | Weeks 1-4 | 1.27 (1.17-1.37) | - | - |
| Dyspepsia | Day 0 | 33.0 (27.4-39.7) | 11.5 (8.82-15.1) | 20.4 (10.9-38.2) |
|  | Weeks 1-4 | 1.58 (1.34-1.88) | 1.11 (0.91-1.35) | 1.07 (0.60-1.89) |
|  | Weeks 5-28 | 1.21 (1.12-1.31) | 1.06 (0.93-1.22) | 1.30 (0.90-1.88) |
|  | Weeks 29-52 | 1.15 (1.06-1.25) | - | - |
|  | Weeks 53-102 | 1.43 (1.25-1.63) | - | - |
| Gallstones | Day 0 | 50.2 (43.1-58.5) | 13.0 (10.4-16.3) | 40.2 (27.6-58.7) |
|  | Weeks 1-4 | 2.96 (2.61-3.35) | 1.28 (1.10-1.49) | 3.74 (2.87-4.88) |
|  | Weeks 5-28 | 1.10 (1.02-1.19) | 0.99 (0.88-1.12) | 1.42 (1.06-1.92) |
|  | Weeks 29-52 | 1.07 (1.00-1.16) | - | - |
|  | Weeks 53-102 | 1.09 (0.96-1.25) | - | - |
| Irritable bowel syndrome | Day 0 | 108 (95.3-121) | 27.5 (22.8-33.1) | 33.9 (19.5-59.0) |
|  | Weeks 1-4 | 2.76 (2.37-3.21) | 1.48 (1.25-1.76) | 2.37 (1.51-3.72) |
|  | Weeks 5-28 | 1.39 (1.28-1.51) | 1.10 (0.96-1.26) | 1.15 (0.73-1.82) |
|  | Weeks 29-52 | 1.20 (1.09-1.32) | - | - |
|  | Weeks 53-102 | 1.55 (1.36-1.78) | - | - |

X insufficient events

#### Supplementary Table 17: Adjusted hazard ratios (95% CI) comparing the incidence of gastrointestinal disease events after COVID-19 with the incidence before or without COVID-19, in the pre-vaccination, vaccinated and unvaccinated cohorts, in individuals without prior history of gastrointestinal operations. Hazard ratios are maximally adjusted.

| Outcome | Time since COVID-19 | Pre-vaccination cohort | Vaccinated cohort | Unvaccinated cohort |
| --- | --- | --- | --- | --- |
| Upper gastrointestinal bleeding | Day 0 | 186 (176-197) | 58.4 (52.5-65.1) | 130 (105-160) |
|  | Weeks 1-4 | 4.09 (3.79-4.41) | 1.89 (1.67-2.14) | 5.09 (4.09-6.35) |
|  | Weeks 5-28 | 1.59 (1.51-1.67) | 1.24 (1.11-1.38) | 1.75 (1.35-2.27) |
|  | Weeks 29-52 | 1.27 (1.20-1.34) | - | - |
|  | Weeks 53-102 | 1.37 (1.25-1.50) | - | - |
| Lower gastrointestinal bleeding | Day 0 | 90.5 (84.6-96.9) | 30.5 (27.0-34.4) | 62.9 (48.8-81.0) |
|  | Weeks 1-4 | 2.52 (2.33-2.73) | 1.40 (1.24-1.57) | 3.30 (2.62-4.17) |
|  | Weeks 5-28 | 1.46 (1.40-1.53) | 1.23 (1.13-1.34) | 1.60 (1.27-2.03) |
|  | Weeks 29-52 | 1.25 (1.20-1.31) | - | - |
|  | Weeks 53-102 | 1.36 (1.25-1.47) | - | - |
| Gastro-oesophageal reflux | Weeks 1-4 | 65.9 (62.5-69.5) | 17.8 (16.2-19.6) | 56.5 (47.6-66.9) |
|  | Weeks 5-28 | 3.23 (3.08-3.39) | 1.39 (1.29-1.49) | 3.87 (3.36-4.45) |
|  | Weeks 29-52 | 1.35 (1.32-1.39) | 1.23 (1.17-1.30) | 1.65 (1.42-1.92) |
|  | Weeks 53-102 | 1.22 (1.18-1.25) | - | - |
|  | Weeks 1-4 | 1.34 (1.28-1.42) | - | - |
| Dyspepsia | Day 0 | 26.9 (24.0-30.3) | 6.18 (4.76-8.02) | 33.9 (25.2-45.4) |
|  | Weeks 1-4 | 1.66 (1.51-1.82) | 1.00 (0.87-1.15) | 2.64 (2.11-3.31) |
|  | Weeks 5-28 | 1.26 (1.21-1.32) | 1.15 (1.05-1.27) | 1.65 (1.33-2.03) |
|  | Weeks 29-52 | 1.24 (1.18-1.29) | - | - |
|  | Weeks 53-102 | 1.17 (1.08-1.28) | - | - |
| Gallstones | Day 0 | 95.7 (88.9-103) | 25.8 (22.7-29.3) | 43.0 (32.2-57.4) |
|  | Weeks 1-4 | 2.71 (2.49-2.96) | 1.33 (1.19-1.50) | 2.96 (2.33-3.75) |
|  | Weeks 5-28 | 1.46 (1.40-1.53) | 1.10 (1.00-1.20) | 1.83 (1.47-2.27) |
|  | Weeks 29-52 | 1.21 (1.15-1.27) | - | - |
|  | Weeks 53-102 | 1.28 (1.18-1.39) | - | - |
| Irritable bowel syndrome | Day 0 | 38.8 (35.0-43.1) | 13.2 (11.3-15.5) | 33.7 (25.7-44.2) |
|  | Weeks 1-4 | 1.92 (1.75-2.11) | 1.12 (0.99-1.27) | 2.95 (2.42-3.60) |
|  | Weeks 5-28 | 1.15 (1.10-1.20) | 1.16 (1.06-1.26) | 1.30 (1.04-1.61) |
|  | Weeks 29-52 | 1.13 (1.08-1.18) | - | - |
|  | Weeks 53-102 | 1.15 (1.05-1.25) | - | - |

X insufficient events

#### Supplementary Table 18. Estimated number of excess diseases for each outcome per 100,000 COVID-19 diagnoses, in the pre-vaccination, vaccinated and unvaccinated cohorts. Increases in risks were estimated within age groups, and the estimated overall increase in risk is the average of these, weighted according to the proportions in each age group in the pre-vaccination cohort.

| Outcome | Pre-vaccination cohort | Vaccinated cohort | Unvaccinated cohort |
| --- | --- | --- | --- |
| Upper gastrointestinal bleeding | 159 | 60 | 112 |
| Lower gastrointestinal bleeding | 144 | 64 | 91 |
| Gastro-oesophageal reflux | 331 | 165 | 296 |
| Dyspepsia | 66 | 29 | 101 |
| Gallstones | 91 | 30 | 106 |
| Irritable bowel syndrome | 66 | 32 | 107 |

### Supplementary Figures

#### Supplementary Figure 1: COVID-19 cases over time

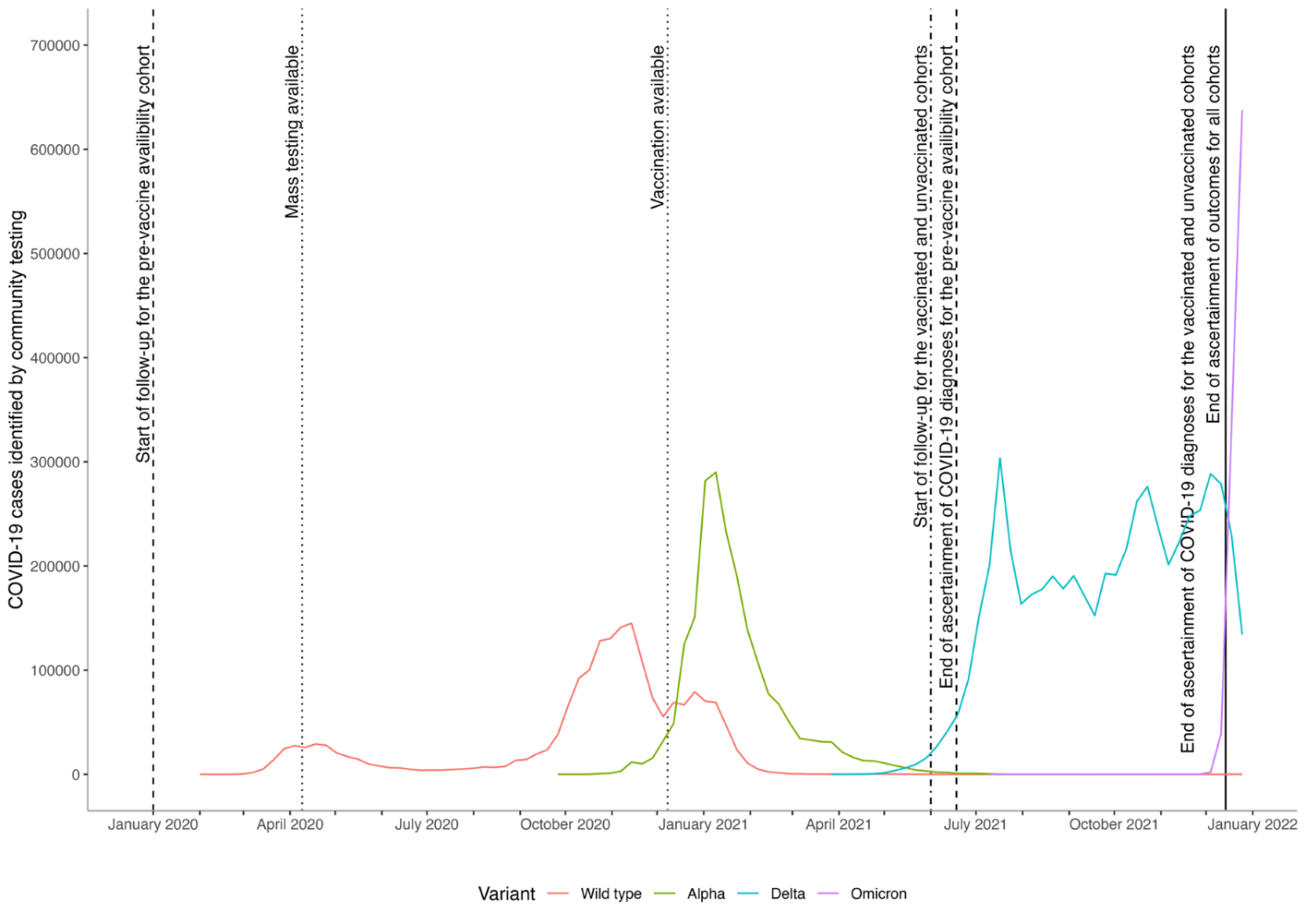

#### Supplementary Figure 2: Diagram showing cohort construction.

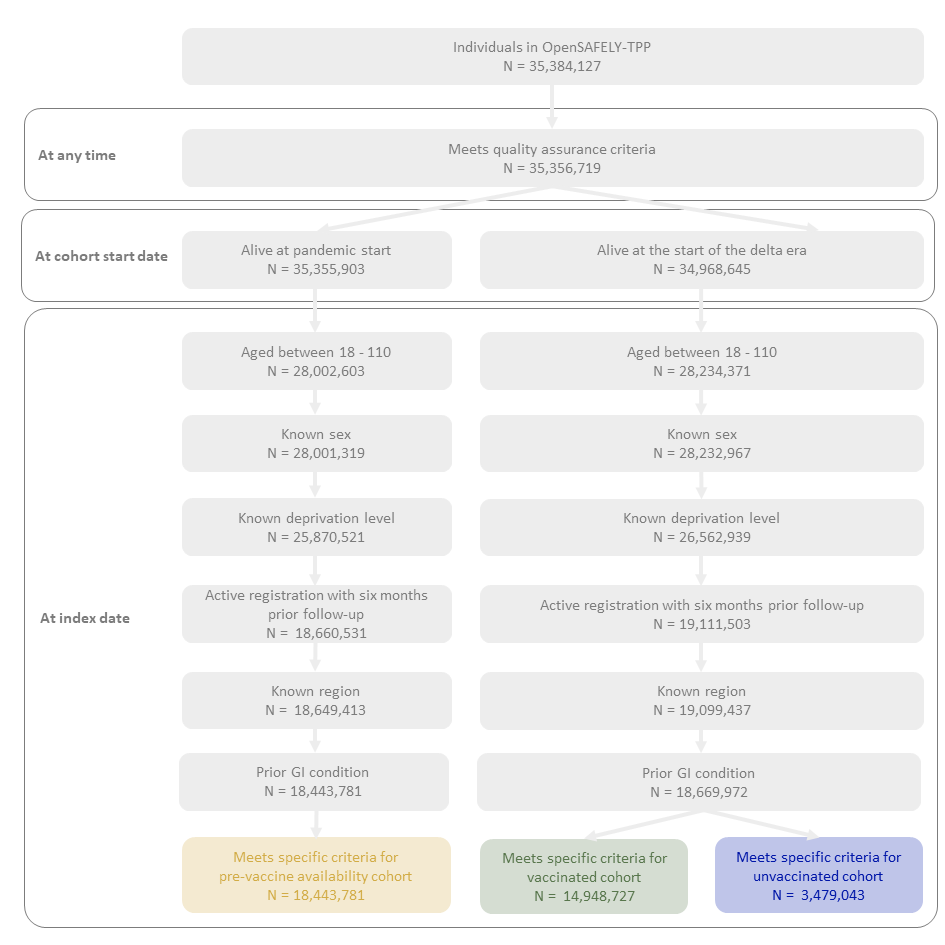

#### Supplementary Figure 3: Maximally adjusted hazard ratios and 95% CIs comparing the incidence of GI outcome diseases after COVID-19 with the incidence before or without COVID-19, in the pre-vaccination, vaccinated and unvaccinated cohorts, by age group. Maximally adjusted hazard ratios

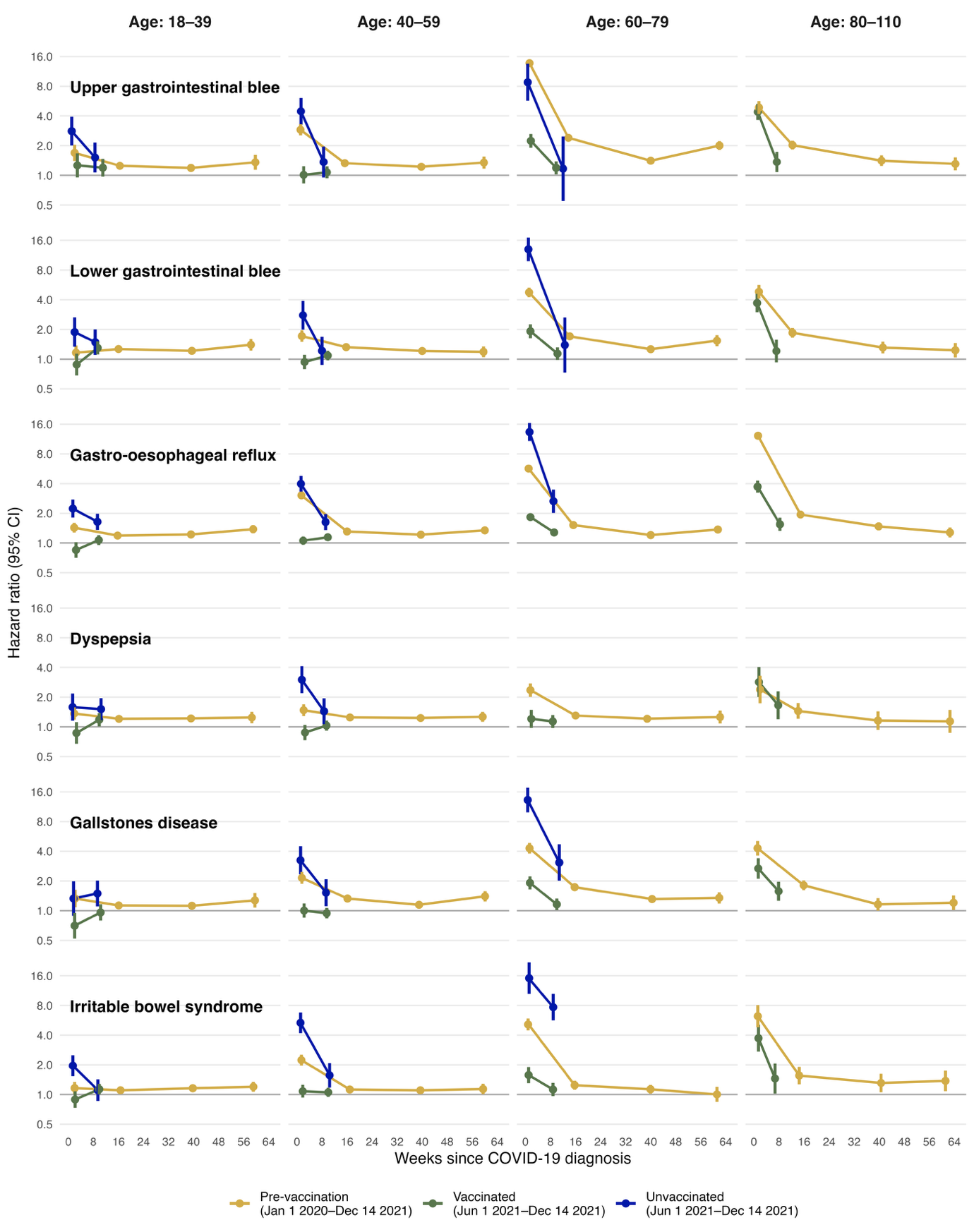

#### Supplementary Figure 4: Maximally adjusted hazard ratios and 95% CIs comparing the incidence of GI outcome diseases after COVID-19 with the incidence before or without COVID-19, in the pre-vaccination, vaccinated and unvaccinated cohorts, by sex. Maximally adjusted hazard ratios

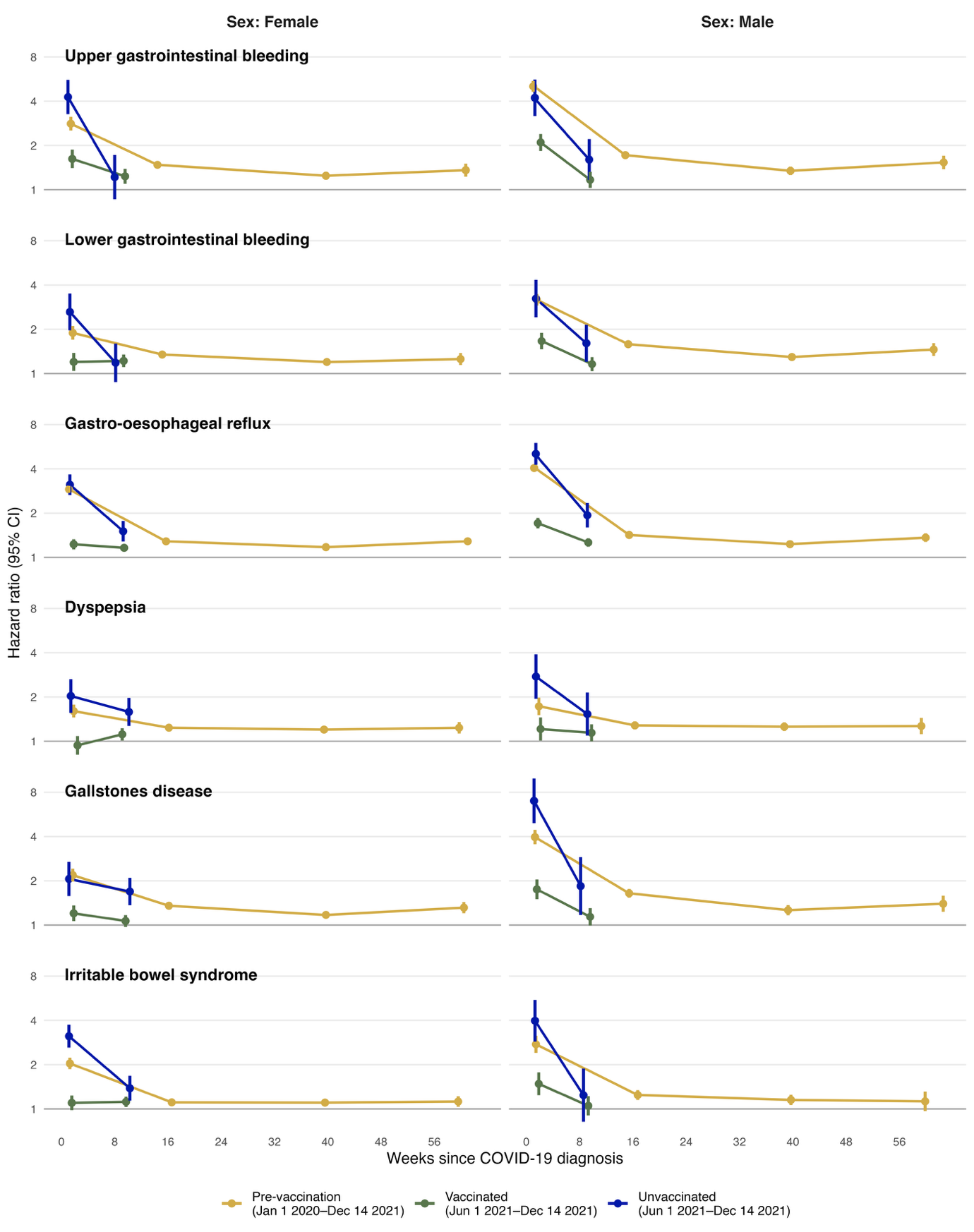

#### Supplementary Figure 5: Maximally adjusted hazard ratios and 95% CIs comparing the incidence of GI outcome diseases after COVID-19 with the incidence before or without COVID-19, in the pre-vaccination, vaccinated and unvaccinated cohorts, by ethnicity. Maximally adjusted hazard ratios

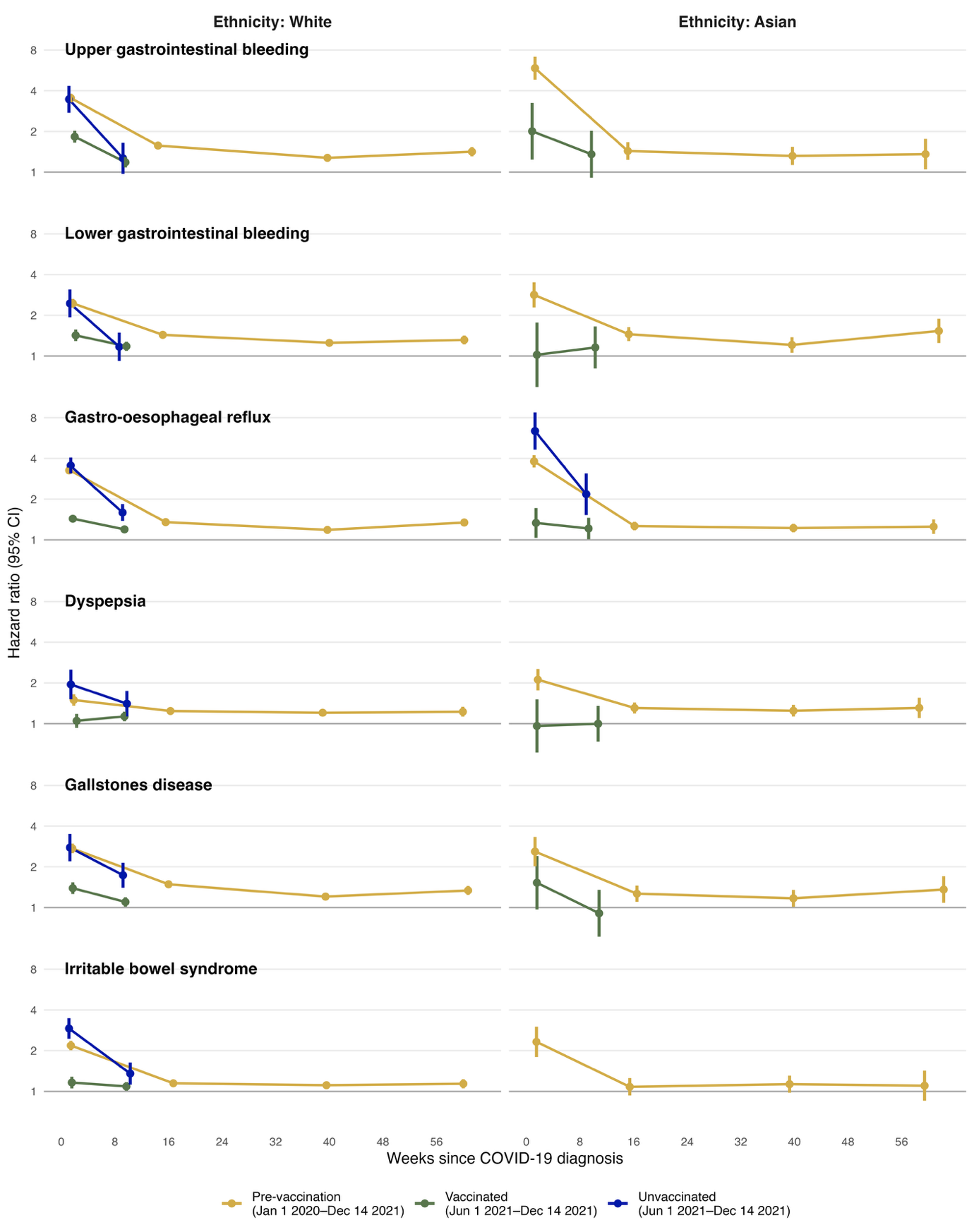

#### Supplementary Figure 6: Maximally adjusted hazard ratios and 95% CIs comparing the incidence of GI outcome diseases after COVID-19 with the incidence before or without COVID-19, in the pre-vaccination, vaccinated and unvaccinated cohorts, by prior history of gastrointestinal operation. Maximally adjusted hazard ratios

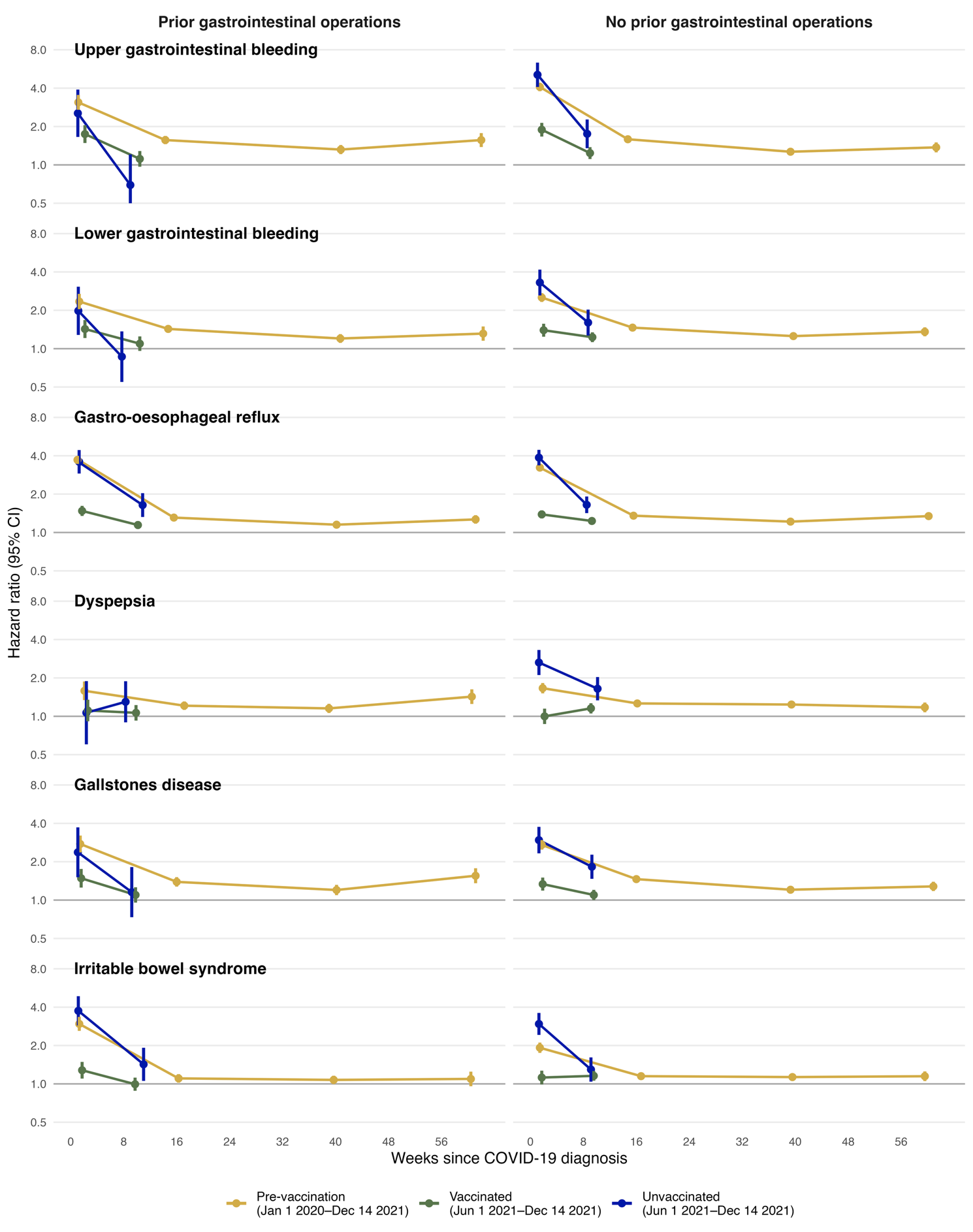

#### Supplementary Figure 7: Maximally adjusted hazard ratios and 95% CIs comparing the incidence of GI outcome diseases after COVID-19 with the incidence before or without COVID-19, in the pre-vaccination, vaccinated and unvaccinated cohorts, by prior history of gastrointestinal event. Maximally adjusted hazard ratios

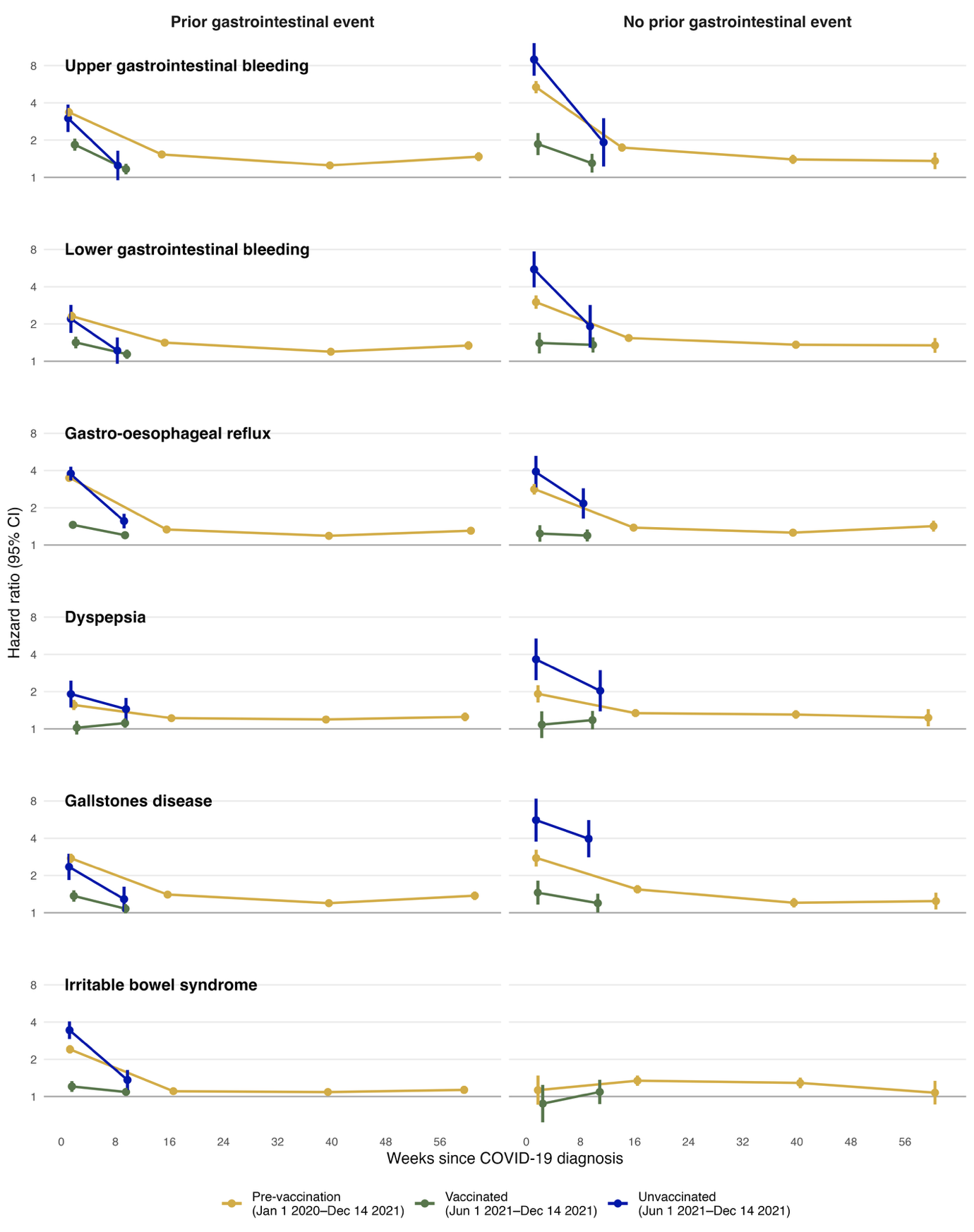

#### Supplementary Figure 8: Estimated absolute increase in risk of upper gastrointestinal bleeding over time since diagnosis of COVID-19, compared with no COVID-19 diagnosis, in the pre-vaccination, vaccinated and unvaccinated cohorts. Increases in risks were estimated within age groups, and the estimated overall increase in risk is the average of these, weighted according to the proportions in each age group in the pre-vaccination cohort.

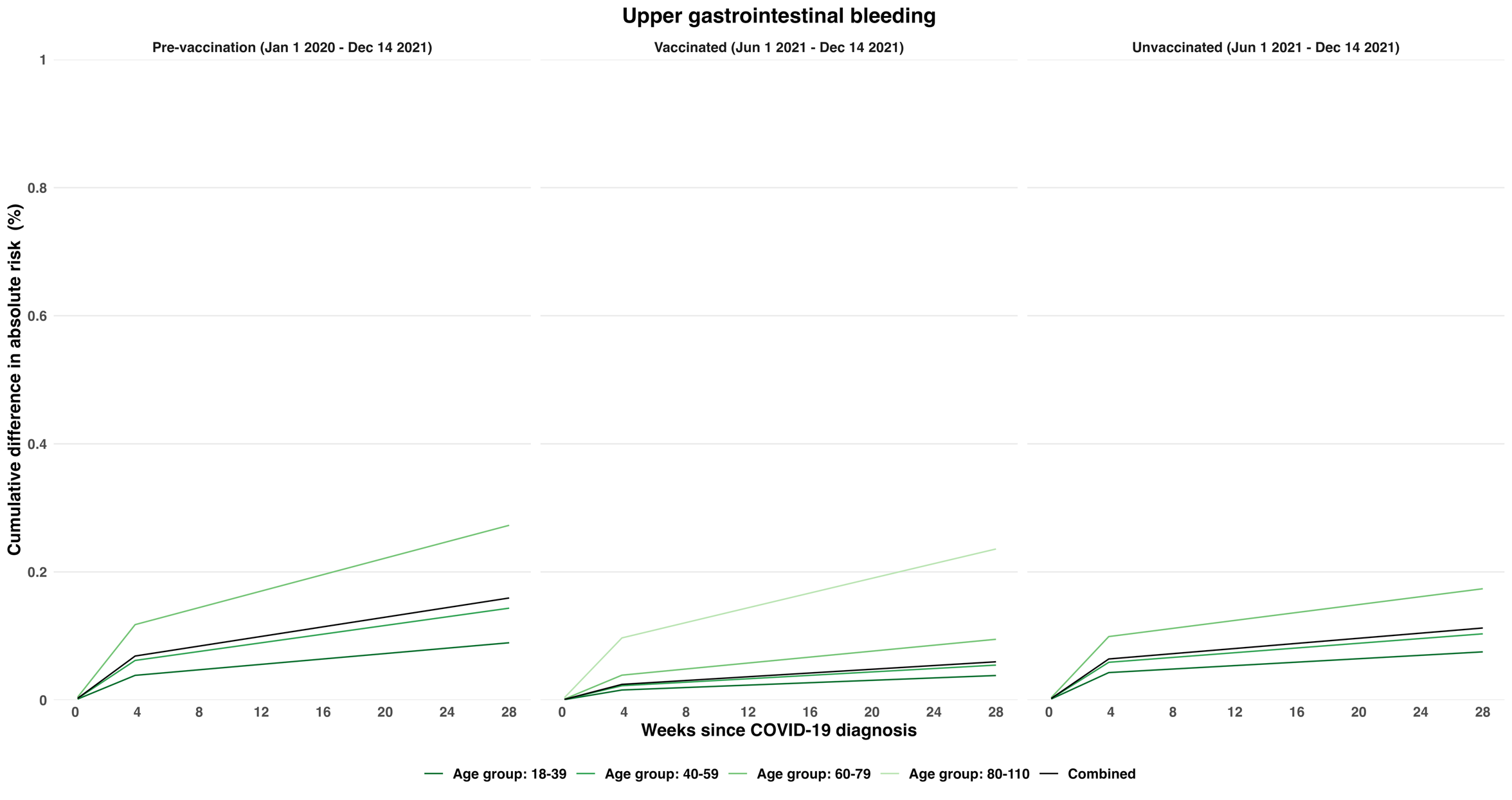

#### Supplementary Figure 9: Estimated absolute increase in risk of lower gastrointestinal bleeding over time since diagnosis of COVID-19, compared with no COVID-19 diagnosis, in the pre-vaccination, vaccinated and unvaccinated cohorts. Increases in risks were estimated within age groups, and the estimated overall increase in risk is the average of these, weighted according to the proportions in each age group in the pre-vaccination cohort.

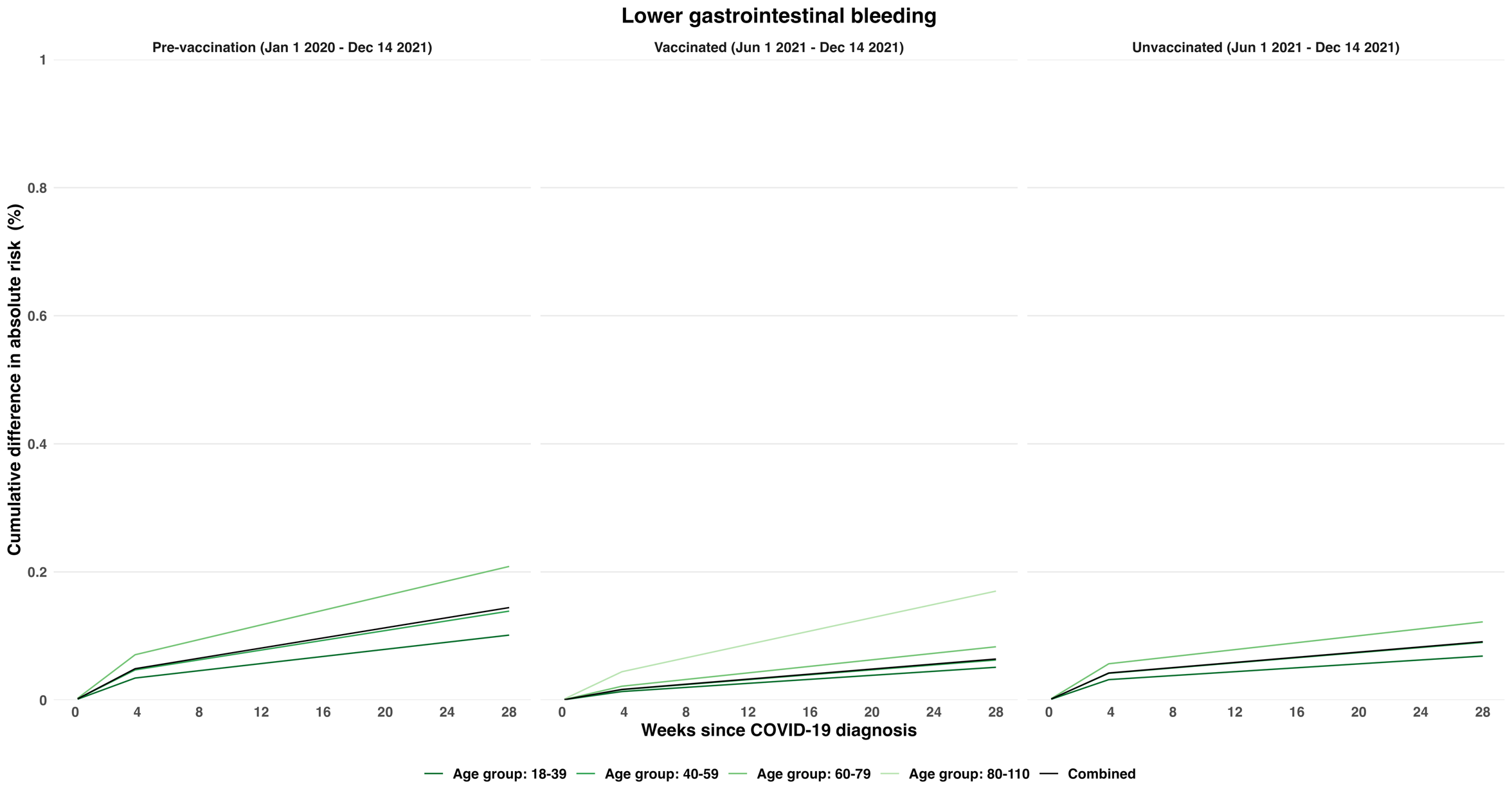

#### Supplementary Figure 10: Estimated absolute increase in risk of gastro-oesophageal reflux over time since diagnosis of COVID-19, compared with no COVID-19 diagnosis, in the pre-vaccination and vaccinated cohorts. Increases in risks were estimated within age groups, and the estimated overall increase in risk is the average of these, weighted according to the proportions in each age group in the pre-vaccination cohort.

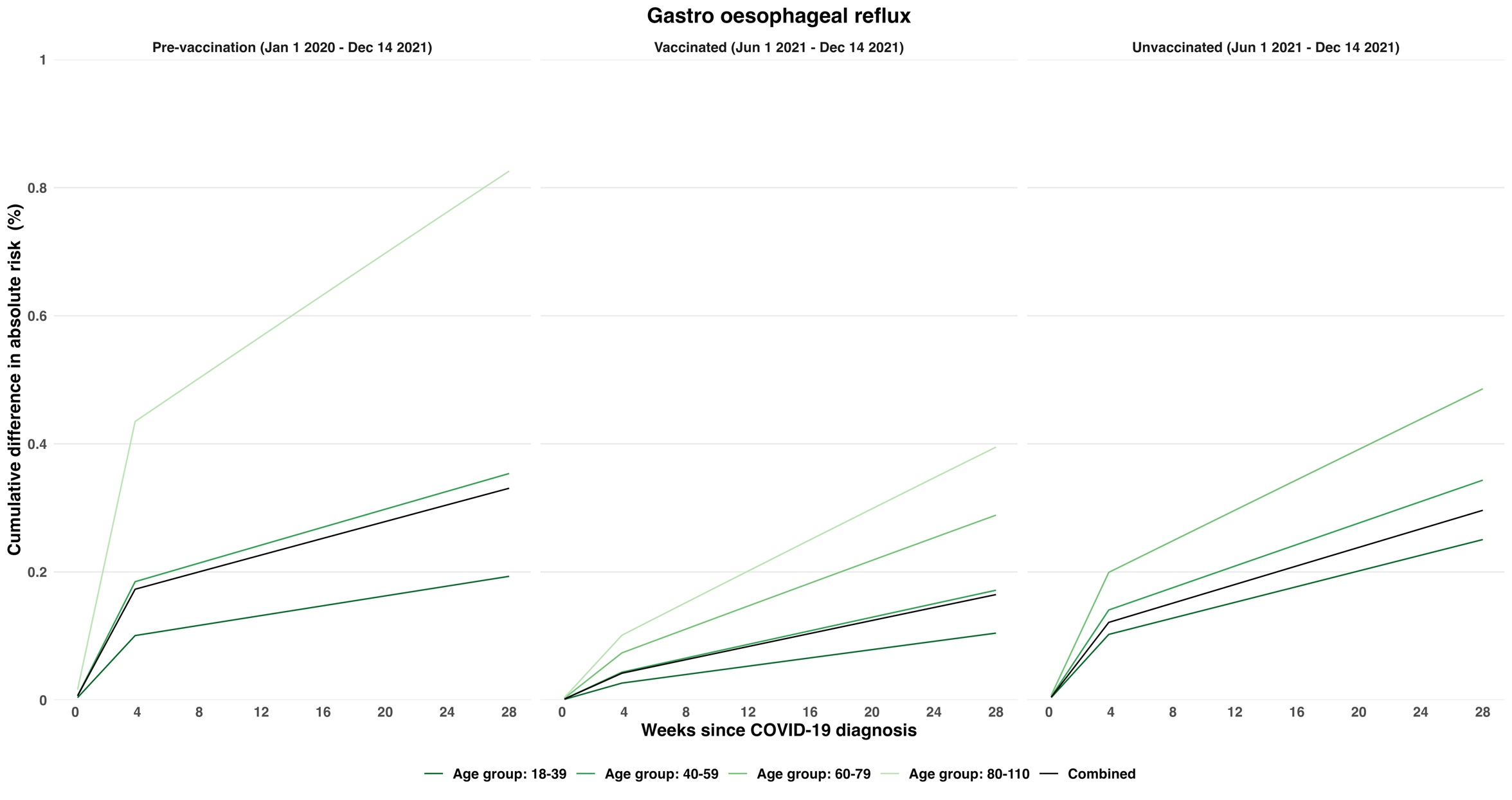

#### Supplementary Figure 11: Estimated absolute increase in risk of dyspepsia over time since diagnosis of COVID-19, compared with no COVID-19 diagnosis, in the pre-vaccination, vaccinated and unvaccinated cohorts. Increases in risks were estimated within age groups, and the estimated overall increase in risk is the average of these, weighted according to the proportions in each age group in the pre-vaccination cohort.

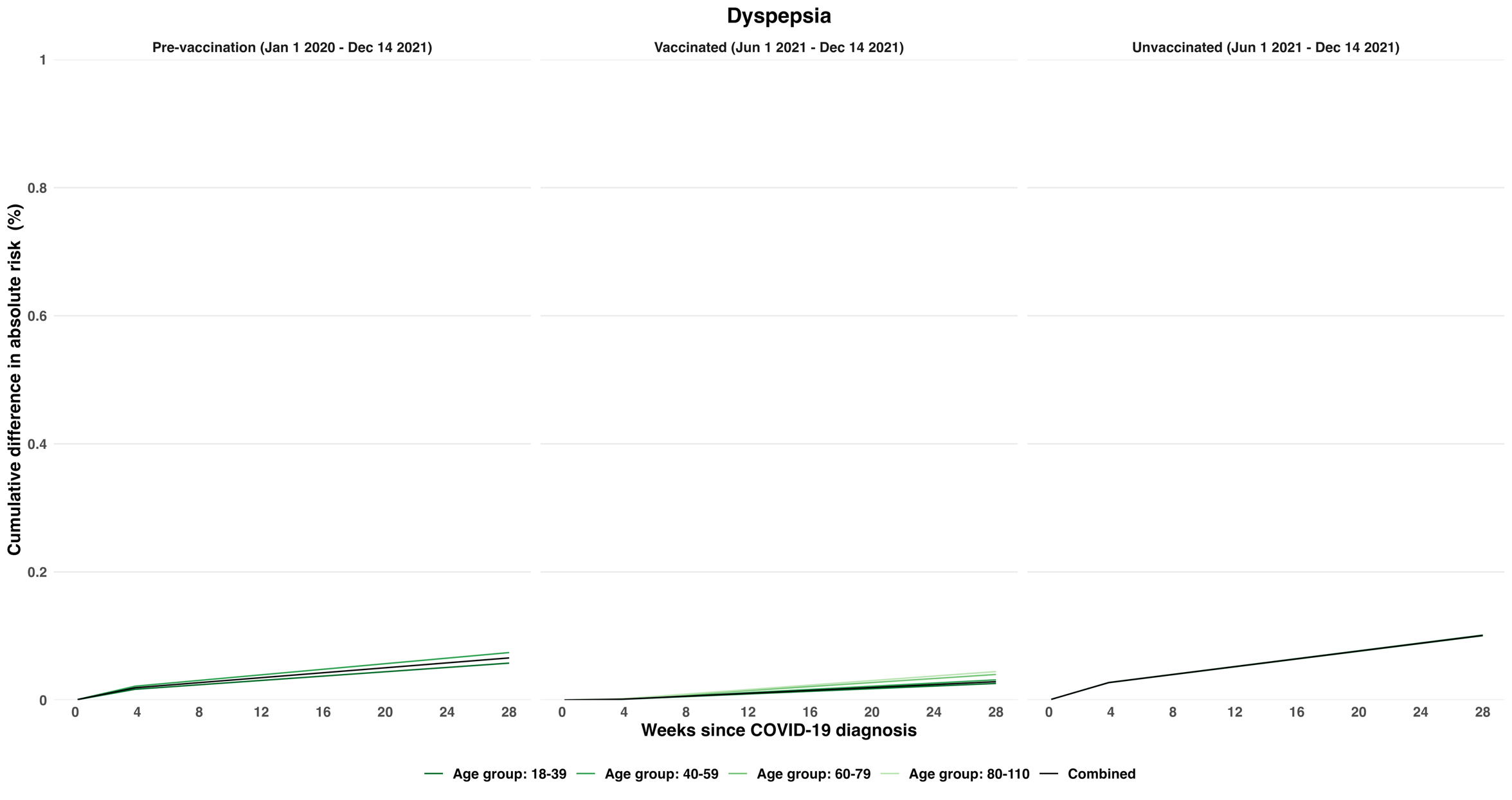

#### Supplementary Figure 12: Estimated absolute increase in risk of gallstones over time since diagnosis of COVID-19, compared with no COVID-19 diagnosis, in the pre-vaccination, vaccinated and unvaccinated cohorts. Increases in risks were estimated within age groups, and the estimated overall increase in risk is the average of these, weighted according to the proportions in each age group in the pre-vaccination cohort.

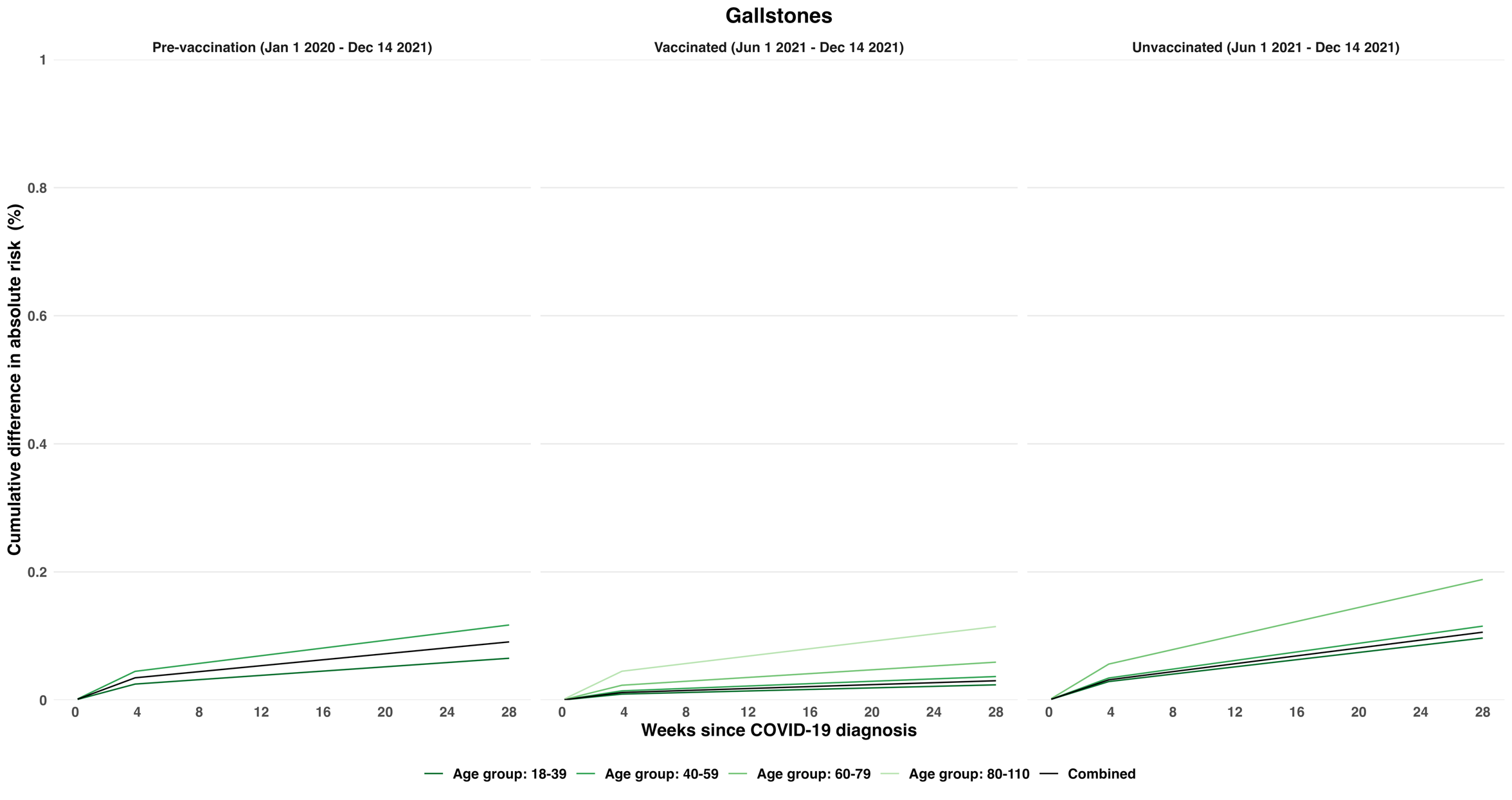

#### Supplementary Figure 13 Estimated absolute increase in risk of irritable bowel syndrome over time since diagnosis of COVID-19, compared with no COVID-19 diagnosis, in the pre-vaccination, vaccinated and unvaccinated cohorts. Increases in risks were estimated within age groups, and the estimated overall increase in risk is the average of these, weighted according to the proportions in each age group in the pre-vaccination cohort.

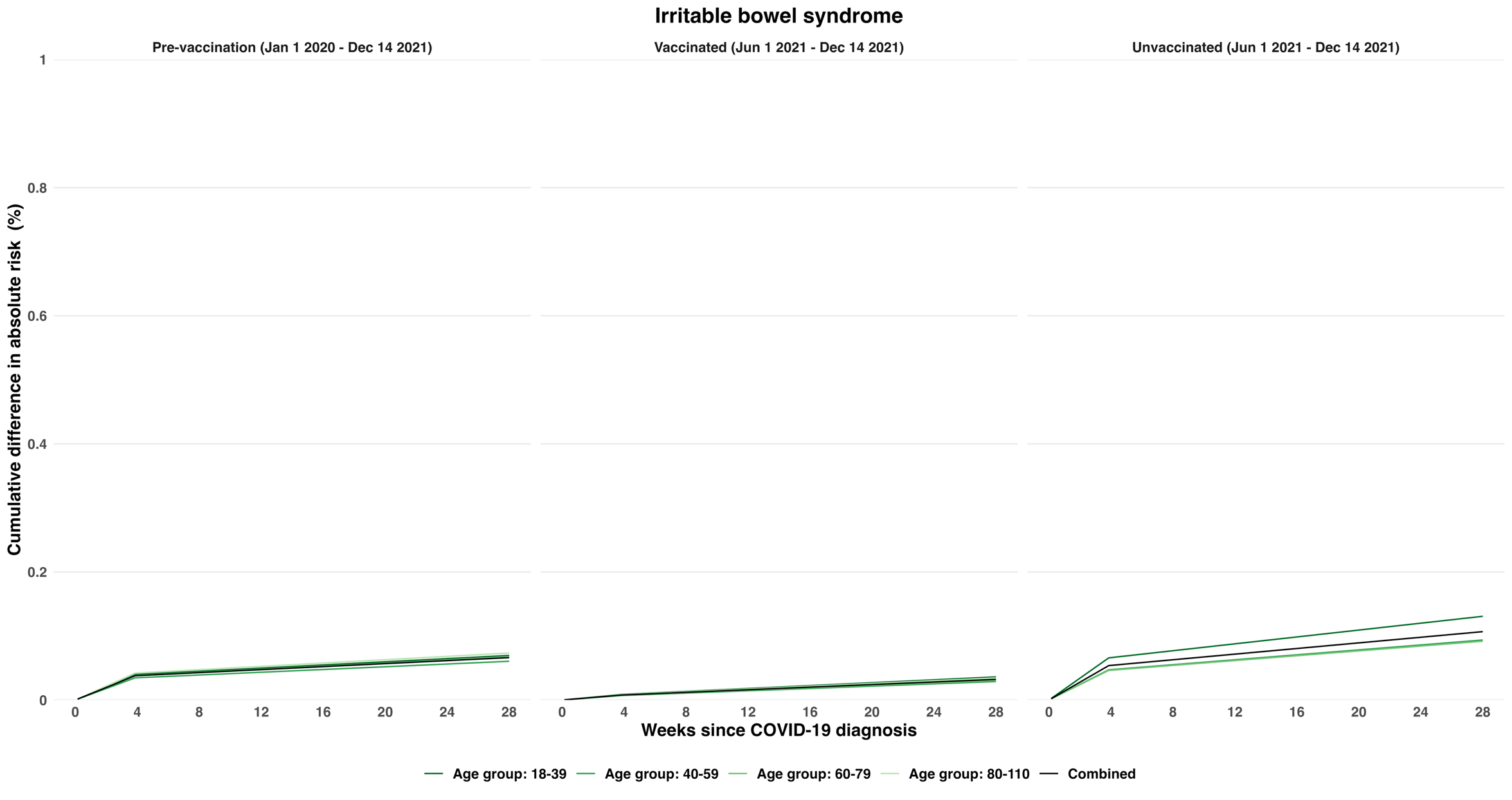
